# Measures of population immunity can predict the dominant clade of influenza A (H3N2) in the 2017-2018 season and reveal age-associated differences in susceptibility and antibody-binding specificity

**DOI:** 10.1101/2023.10.26.23297569

**Authors:** Kangchon Kim, Marcos C. Vieira, Sigrid Gouma, Madison E. Weirick, Scott E. Hensley, Sarah Cobey

**Author notes:** These authors contributed equally (Kangchon Kim, Marcos C. Vieira).

## Abstract

**Background:** For antigenically variable pathogens such as influenza, strain fitness is partly determined by the relative availability of hosts susceptible to infection with that strain compared to others. Antibodies to the hemagglutinin (HA) and neuraminidase (NA) confer substantial protection against influenza infection. We asked if a cross-sectional antibody-derived estimate of population susceptibility to different clades of influenza A (H3N2) could predict the success of clades in the following season.

**Methods:** We collected sera from 483 healthy individuals aged 1 to 90 years in the summer of 2017 and analyzed neutralizing responses to the HA and NA of representative strains using Focus Reduction Neutralization Tests (FNRT) and Enzyme-Linked Lectin Assays (ELLA). We estimated relative population-average and age-specific susceptibilities to circulating viral clades and compared those estimates to changes in clade frequencies in the following 2017-18 season.

**Results:** The clade to which neutralizing antibody titers were lowest, indicating greater population susceptibility, dominated the next season. Titer correlations between viral strains varied by age, suggesting age-associated differences in epitope targeting driven by shared past exposures. Yet substantial unexplained variation remains within age groups.

**Conclusions:** This study indicates how representative measures of population immunity might improve evolutionary forecasts and inform selective pressures on influenza.

## Introduction

The epidemiological and evolutionary dynamics of antigenically variable pathogens are intrinsically sensitive to immunity in the host population. This understanding has long shaped vaccination strategies against influenza. Twice each year, representative strains from circulating clades are evaluated for their ability to escape antibodies to current vaccine strains, under the expectation that these clades might come to dominate and could be poorly matched by the current vaccine. As surrogates for the human population, influenza-naive ferrets are infected or vaccinated with one of a set of reference influenza strains (e.g., current vaccine strains), and their post-exposure sera are tested against candidate strains for the next vaccine. The extent to which these sera cross-react or neutralize candidate strains is taken as a measure of their immune escape or antigenic distance [1, 2]. These experimental measures of immune escape, alongside other estimates of variant growth rates and sequence-based fitness models [3], are used to anticipate the dominant clade and need for vaccine updates. In the past few years, escape from human sera has been considered too (e.g., [4]).

An open question is whether more direct and representative estimates of population immunity could lead to better vaccine choices while potentially shedding light on the mechanisms of coevolution between the viral population and host immunity. In the past decade, large differences have occasionally appeared in the antigenic distances inferred from ferret compared to human sera [5, 6]. These differences might arise at the species level, although the antibody responses of ferrets and humans after their first influenza exposures appear roughly similar [7]. A more likely explanation comes from observations of original antigenic sin, whereby individuals exposed to the same strain of influenza can mount antibody responses with different cross-reactivity profiles shaped by their distinct histories of exposure [5, 8–11]. These past infections and vaccinations lead to biases in which viral sites or epitopes antibodies recognize. Consequently, a mutation in one epitope might be antigenically important for some people (or ferrets) but not others. Since most influenza infections occur in people with preexisting immunity to influenza, and antibodies to influenza surface proteins contribute substantially to protection and transmission [5, 12–16], accurate measures of population immunity may be useful in viral forecasting and vaccine strain selection.

Using the 2017-2018 influenza season in North America as a case study, we characterized a cross-sectional, age-representative estimate of antibody-mediated immunity in an urban population and asked whether it could predict which of several circulating clades of H3N2 would dominate regionally in the next influenza season. Forecasting for vaccine strain selection often focuses on antigenic changes to the hemagglutinin (HA) surface protein, which vaccines attempt to match. We measured neutralizing antibody titers to the neuraminidase (NA) protein as well as to HA because antibodies to NA are also protective and should thus affect clade fitness. We found large differences in the expected susceptibility of the population to different clades’ HA and NA, and these differences in susceptibility predicted clade dominance. They also partially predicted the relative attack rates of clades by age. We furthermore quantified the heterogeneity in neutralizing titers in the population, finding patterns consistent with age-associated epitope targeting. Although data from a single timepoint cannot fully elucidate the role of population immunity in clade evolution, our results demonstrate for the first time how such measures can improve on traditional approaches.

## Results

### Human sera from the summer of 2017 poorly neutralize the clade that dominated in North America in the next influenza season

We investigated whether neutralizing antibody titers to HA and NA from H3N2 clades circulating in early 2017 could predict the dominant (most frequent) clade in the next influenza season. Antibodies to HA can protect against infection [12, 13, 15–17], and we expected that the clade to which the largest fraction of the population had poorly neutralizing anti-HA titers would be most successful. This expectation implicitly assumes that exposure rates, other factors affecting susceptibility, and the average infectiousness or transmissibility of an infected person do not differ starkly between age groups; it also assumes that antibody-mediated protection derives primarily from neutralization and not Fc-mediated effector functions, or that the two are well correlated.

Antibody neutralization was measured by the focus reduction neutralization test (FRNT) for anti-hemagglutinin antibodies and enzyme-linked lectin assay (ELLA) for the anti-neuraminidase antibodies, and these antibodies levels were assumed proxies for protection. Correlates of protection have not been established for FRNT-derived titers, but because microneutralization titers correlate well with hemagglutination inhibition (HAI) [18], and a 1:40 HAI titer is traditionally associated with a 50% reduction in infection risk [12], we initially assumed a 1:40 FRNT titer corresponds to a 50% chance of infection, testing other assumptions in sensitivity analyses. We looked at the fraction of the population below this cutoff for each clade to obtain the expected relative susceptibility and ranked clades by this measure. Using a cutoff avoids overestimating protection that might arise from especially high titers in a subset of recently infected individuals, but for robustness, we also estimated the relative susceptibility according to the geometric mean titer (GMT) to each clade, with lower GMT implying higher susceptibility. With both measures, the population-level susceptibility was estimated by weighting the susceptibility of different age groups according to their proportion in the population (Methods). Protective thresholds for ELLA NA titers have not yet been established. We initially assumed 1:80 to be the 50% protective titer and later explored other assumptions.

We collected serum samples from May to August of 2017 from the University of Pennsylvania BioBank and Children’s Hospital of Philadelphia [19] (Methods; Fig S1). Samples from children were primarily obtained for lead testing. Adults with certain health conditions were excluded. Since we knew the age of each serum donor, we were able to adjust our estimates of population immunity to reflect the age distribution of the United States population (Methods). However, no information on vaccination status was available, and therefore we could not adjust our estimates to reflect vaccination status in the general population. We measured neutralizing titers to the 8 HA and 2 NA representing common current or recently circulating H3N2 clades (Fig 1A left for HA and Fig S2A left for NA).

**Fig 1.**
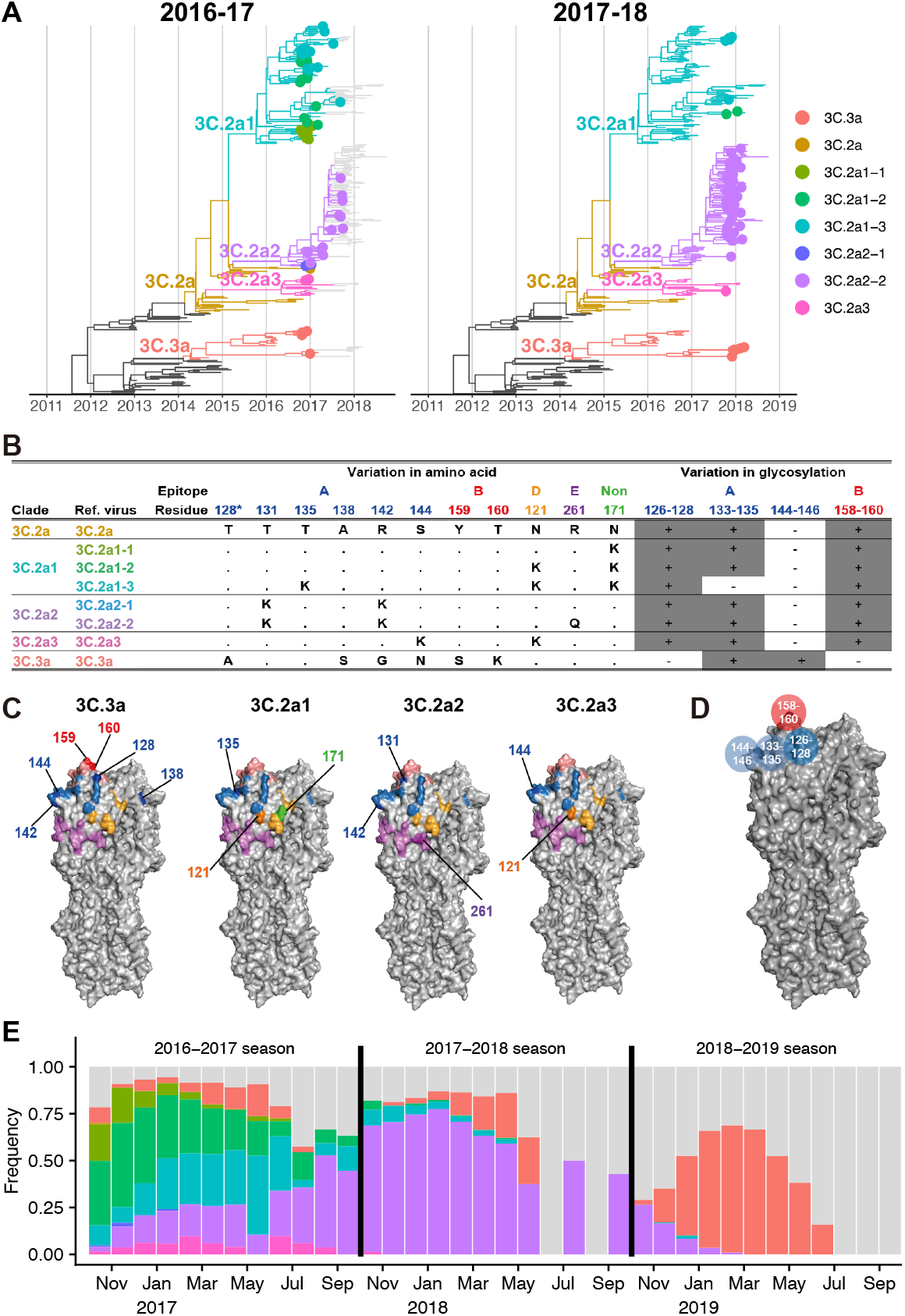
Reference viruses representing co-circulating H3N2 clades during the 2016-2017 season. A. Genealogies of H3N2 HA through the 2016-2017 (left) and 2017-2018 season (right). Tips are colored by similarity to reference viruses. For clades with multiple representative viruses, branches are colored the same as one of those viruses, chosen arbitrarily. Tips are shown as filled circles if collected in North America during the most recent season. B. Amino acid and glycosylation site variation among reference viruses. Clades 3C.3a and 3C.2a diverge at additional non-epitope sites (not shown). Residue 128 belongs to antigenic site B, but the substitution T128A results in loss of glycosylation on residue 126 of epitope A. Therefore, we show residue 128 in epitope A and in the glycosylation site involving residues 126-128, following [1]. C. Variable residues among the reference viruses are shown on the H3 structure of A/Aichi/2/1968 (Protein Data Bank: 1HGG) and colored by epitope as in panel B. For each strain, residues differing from 3C.2a are numbered and darker in color. D. Glycosylation sites used in the model shown on the H3 structure. E. Monthly clade frequencies among GISAID isolates from North America collected between the 2016-2017 and the 2018-2019 seasons. Gray bars represent isolates that do not belong to the focal clades in this study.

To represent the circulating diversity of H3N2 viruses, we identified major circulating clades by inspecting the nextstrain [20] H3N2 genealogy in the early summer of 2017 (note that the genealogy in Fig 1A was constructed later with data up to January 2022). Two distinct clades, 3C.2a and 3C.3a, which last shared a common ancestor in 2012, circulated globally. These clades differed by amino acid substitutions in epitopes A and B (Fig 1B, C) and in non-epitope sites. Clade 3C.2a had gained a potential glycosylation site at epitope B (K160T; H3 numbering used throughout) and had lost a glycosylation motif at epitope A (N144S). Clade 3C.3a had lost a different glycosylation site in epitope A (T128A) (Fig 1B, C). We chose the vaccine strain A/Hong Kong/4801/2014 to represent clade 3C.2a and the vaccine strain A/Switzerland/9715293/2013 to represent 3C.3a, both close to the root of their respective clades.

In addition to those two major clades, we further split clade 3C.2a into subclades 3C.2a1, 3C.2a2, and 3C.2a3. We constructed representative sequences for these subclades by introducing mutations into the sequence of the 3C.2a reference strain. For subclades 3C.2a1, 3C.2a2, and 3C.2a3, we constructed 3, 2, and 1 reference viruses, respectively, each carrying subclade-specific nonsynonymous substitutions and (for 3C.2a1 and 3C.2a2) potentially important amino acid polymorphisms within the subclade. Each subclade contained an epitope A substitution compared to the 3C.2a reference strain (Fig 1B, C). Notably, one reference virus for clade 3C.2a1 (virus 3C.2a1-3) had the T135K mutation, which removes a glycosylation motif in epitope A. Except for 3C.2a1-3, all representative strains had identical sequences to one or more naturally occurring strains (Table S1). Only the 3c.2a reference virus was represented among candidate vaccine viruses for the 2017-2018 season (identical to A/Hong Kong/4801/2014, A/Hawaii/47/2014, A/Victoria/673/2014 and A/Norway/2178/2014). Once we had determined the sequences of the representative strains, we constructed the viruses by reverse genetics (Methods). We used the same set of reference viruses for all serum donors.

For all reference viruses, an undetectable HA titer (titer of 1:10) was the most common HA titer in all age groups except children 5-17 years old (Figs 2A, S3). Most people over 4 years old had detectable NA titers (1:≥ 20) (Figs 3A, S4). Even though detectable antibody to H3N2 HA or NA is expected among older children and adults, who have been infected and possibly vaccinated with H3N2, surprisingly large variation was observed among individuals of the same age (Figs 2A, 3A). These are likely genuine differences in titer, as technical replicates had high agreement.

**Fig 2.**
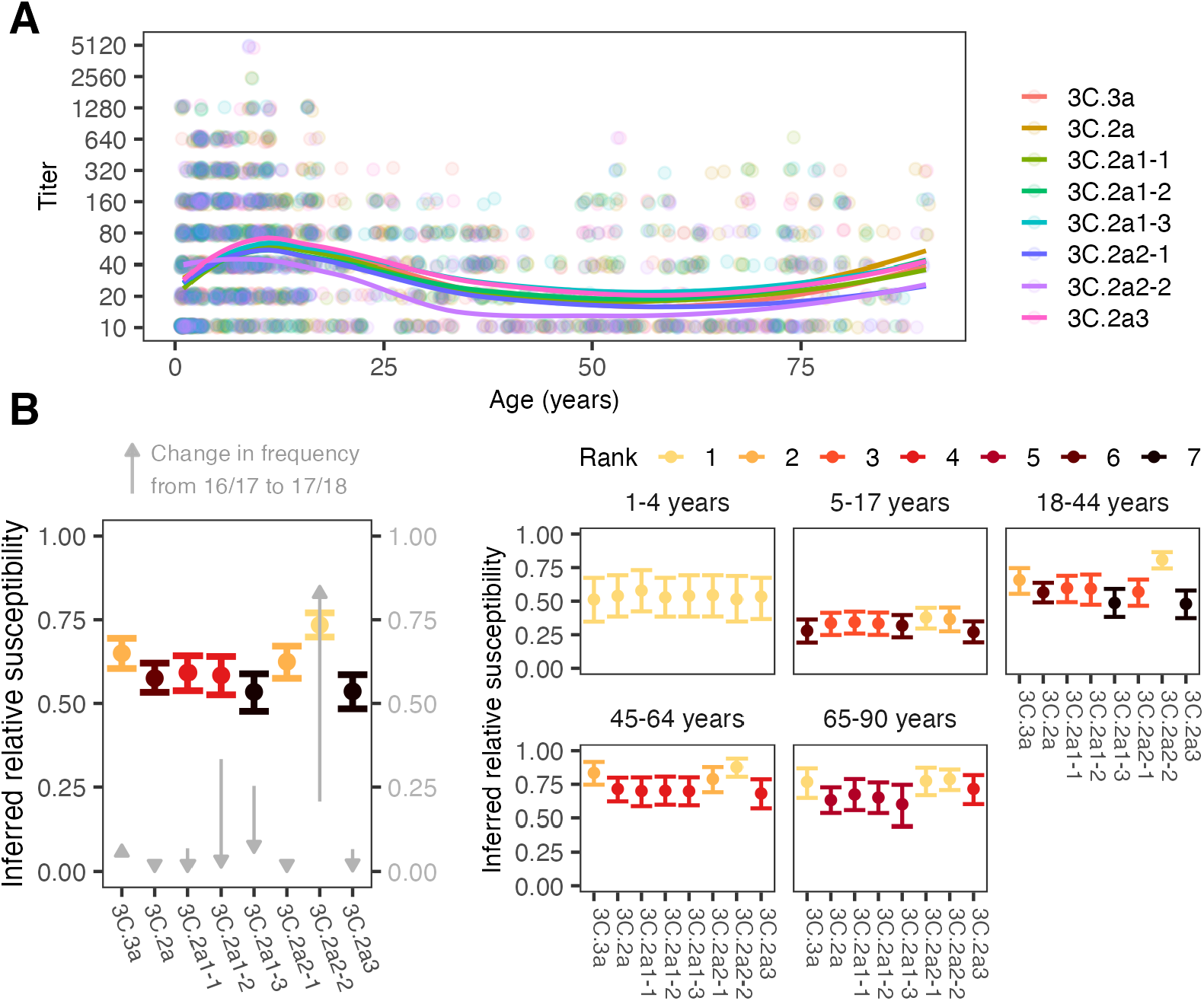
Antibody titers and inferred relative susceptibilities to co-circulating H3 strains show variability by strain and age group. A. FRNT titers with points jittered slightly along the x- and y-axes. Lines are locally estimated scatterplot smoothing (LOESS) curves of geometric mean titers (smoothing parameter *α* = 0.75, degree = 2). B. Inferred relative susceptibility to each reference strain for the whole population (left) and by age group (right). The bars indicate 95% CIs obtained from bootstrapping. Separately for the overall population and each age group, we ranked strains in decreasing order of susceptibility using pairwise bootstrap tests (e.g., a strain had rank 1 if susceptibility to it was significantly higher than susceptibility to all other strains in pairwise tests, 2 if significantly higher for all but one strain, etc. Strains are tied in rank and appear in the same color if their relative susceptibilities do not differ significantly. The gray arrows show how the clades changed in frequency in North America between the 2016/17 and 17/18 seasons.

**Fig 3.**
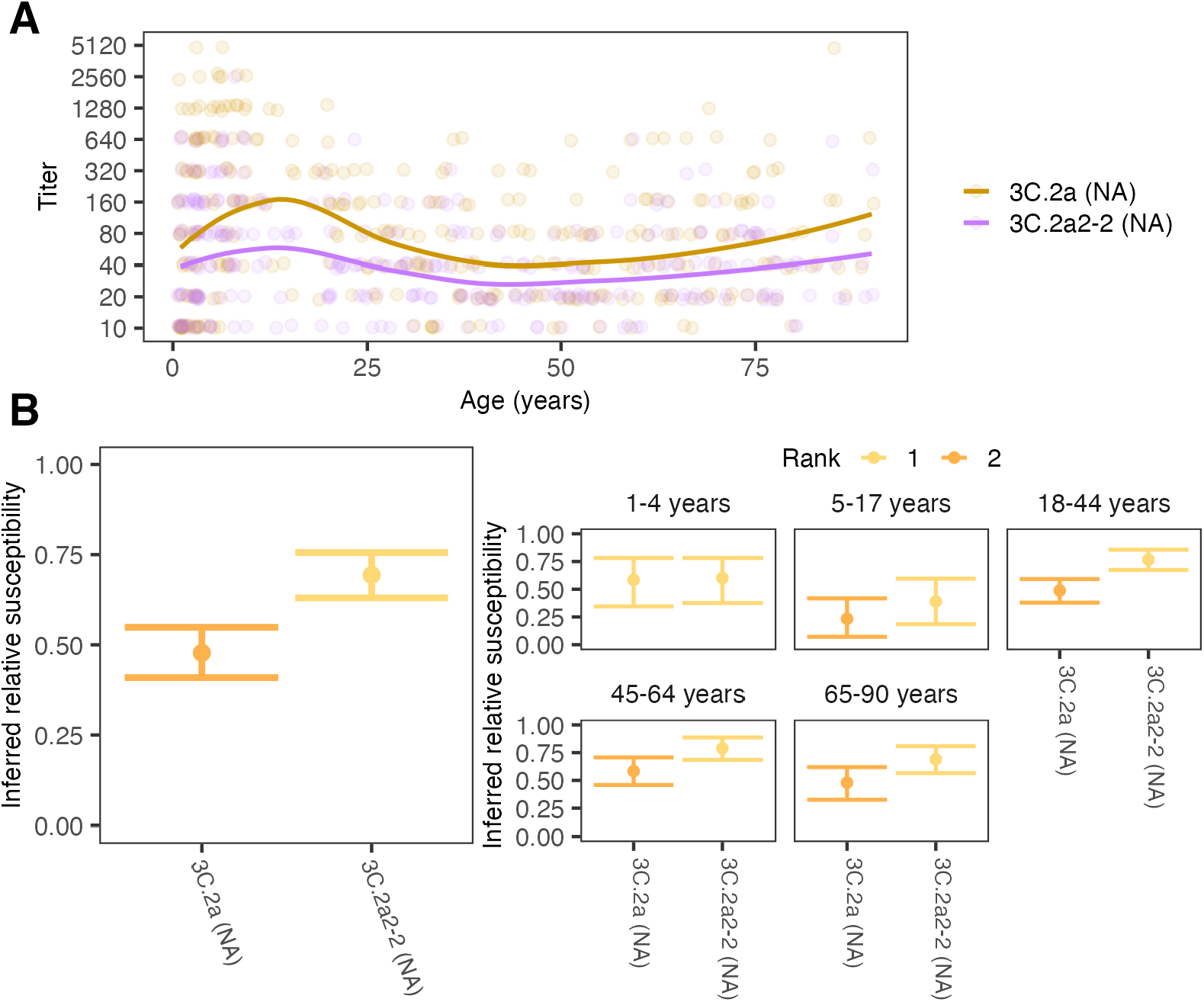
Antibody titers and relative susceptibilities to co-circulating N2 strains show differences by strain and age group. A. ELLA titers with points jittered slightly along the x- and y-axes. Lines are locally estimated scatterplot smoothing (LOESS) curves of geometric mean titers (smoothing parameter *α* = 0.75, degree = 2). B. Inferred relative susceptibility and its rank for each NA for the whole population (left) and by age group (right). A lower rank indicates significantly higher susceptibility. Strains are tied in rank and appear in the same color if their relative susceptibilities do not differ significantly.

The population-level relative susceptibility inferred using the 1:40 protective cutoff in HA titer was highest to the 3C.2a2 subclade, specifically the group of viruses with 261Q in epitope E (3C.2a2-2 reference strain; the susceptibility to 3C.2a2-2 is higher than the susceptibility to 3C.2a2-1 and 3C.3a, both bootstrap *p <* 0.001), followed by the rest of the 3C.2a2 subclade (3C.2a2-1 reference strain; the susceptibility to 3C.2a2-1 is higher than the susceptibility to 3C.2a1-1, bootstrap *p <* 0.05) and the 3C.3a clade (*p <* 0.01 for the same test; Fig 2B, left panel) (Methods). Not only was the average population susceptibility highest to 3C.2a2-2, but susceptibility to it was also the highest or the second highest among the reference strains for all age groups except 1-4 year-olds. Using GMTs or alternative titer cutoffs also suggested the susceptibility was highest to the 3C.2a2-2 reference strain followed by 3C.2a2-1 (Figs S5-S8). The greatest protection or lowest susceptibility in the population by both measures was to strains of the 3C.2a1 subclade with 135K in epitope A and 121K in epitope D (reference strain 3C.2a1-3) and subclade 3C.2a3 (reference strain 3C.2a3).

Consistent with simple predictions, clade 3C.2a2 dominated in North America in the 2017-18 season (Fig 1A, right panel; Fig 1E; Fig 2B, gray arrows), followed by 3C.3a. To assess dominance, influenza sequences were downloaded from GISAID [21]. We assigned 9913 sequences collected in North America during the 2016-2017 and 2017-2018 influenza seasons to reference viruses based on their genetic similarity at segregating sites and found that the frequency of sequences genetically similar to reference strain 3C.2a2-2 in clade 3C.2a2 increased from 21% in the 2016-2017 season to 85% in the 2017-2018 season (Fig 2B; Fig S9). Clade 3C.3a increased from 6% to 8% over that period. We did not find a perfect correlation between the rank measured by inferred relative susceptibilities and rank by relative growth: despite having higher estimated susceptibility than subclade 3C.2a1 (3C.2a1-3), subclade 3C.2a1 (3C.2a1-2) experienced a more severe decline. Although the available sequences are not generated from any kind of systematic surveillance program and thus may not accurately reflect relative prevalence, trends were stable regionally (Fig S9A). The results suggest that population-average anti-HA neutralizing titers reflect strain fitness, but that other factors may be relevant for detailed predictions.

We next measured antibody responses to NA reference strains representing the NAs of clades 3C.2a and 3C.2a2 (“3C.2a (NA)” and “3C.2a2-2 (NA)”, respectively) (Fig 3A) [19]. The two reference viruses differ by 7 amino acid substitutions in the NA head: 176, 245, 247, 329, 334, 339, and 386. We first estimated population-level relative susceptibilities to the two clades using a 1:80 protective cutoff (Fig 3B, left panel).

Similar to our findings for HA, serological responses to NA indicated higher susceptibility to 3C.2a2-2 (NA) than to 3C.2a (NA) across all age groups, consistent with a positive correlation between HA and NA titers across all age groups and especially in children (Fig S10). Using GMT or alternative NA titer thresholds also suggested higher susceptibility to 3C.2a2-2 (Figs S11-S13). Because only two NA reference strains were used, we cannot conclude if anti-NA titers would have predicted clades’ rank frequencies as accurately or perhaps better than titers to HA, but they are generally consistent with higher susceptibility to the 3C.2a2 clade compared to the ancestral 3C.2a.

### Age groups differ in their susceptibility to and relative attack rates with different H3N2 clades

Because age-specific patterns of antibody titers have been associated with age-specific infection risk [5, 22], we estimated relative susceptibility to each clade within each age group and measured correlations with their estimated relative clade-specific infection rates in the 2017-2018 influenza season. Age groups differed slightly in their expected susceptibilities to different clades of H3N2 (Fig 2B, right panel). Assessed by their anti-HA titers, children 1 to 4 years old appear equally susceptible to all reference viruses. The anti-HA titers of older children and adults showed heightened susceptibility to the 3C.2a2 clade: titers from 5-to 17-year-olds indicated the highest susceptibility to the basal 3C.2a2 clade (reference strain 3C.2a2-1) followed closely by reference strain 3C.2a2-2, the 3C.2a2 subclade with the R261Q substitution. People aged 18-64y had pronounced susceptibility to reference strain 3C.2a2-2 compared to other clades.

Because 3C.2a2-2 differs from 3C.2a2-1 by a single amino acid substitution (R261Q), these results suggest that many HA antibodies in adults target the mutated site. All age groups with previous influenza experience (≥ 5 y) were least susceptible to clades 3C.2a1 and 3C.3a (reference strains 3C.2a1-3 and 3C.2a3, respectively). Interestingly, 5-to 17-year-olds were least susceptible to 3C.3a, while adults were relatively susceptible to 3C.3a. We also found that children 1 to 4 years old had comparable susceptibility to the two clades of NA, and all older age groups demonstrated greater susceptibility to the 3C.2a2 clade (3C.2a2-2 (NA)) (Fig 3B, right panel).

We evaluated whether the age-associated trends in relative susceptibilities to different clades in the summer of 2017 were mirrored in their relative rate of infection with each clade in the 2017-2018 influenza season. Due to lack of systematic surveillance, unbiased estimates of attack rates by clade do not exist for this population. We nonetheless examined the ages associated with sequences uploaded into GISAID to approximate the proportion of infections caused by each clade in each age group. Because the 3C.2a2 clade dominated in the 2017-2018 season and all but the youngest age groups showed particularly high susceptibility to this clade, we expected clade 3C.2a2 to be the most frequent within each age group. This is what we found (Fig 4, Figs S14-S15). However, we observed that children *<* 5 y old, who seemed approximately equally susceptible to all clades by HA and NA, had a relatively lower proportion of 3C.2a2 infections compared to adults (chi-square test, *p <* 0.001). Children 5-17 y old, who were only slightly more susceptible to 3C.2a2 than other clades, also had a lower proportion of 3C.2a2 infections compared to adults (*p <* 0.001). Consistent with our observation that 18-to 64-year-olds were disproportionately susceptible to clade 3C.2a2, the age distribution of that clade was slightly more skewed toward adults compared to non-3C.2a2 clades, which were more common in children (Fig 4).

**Fig 4.**
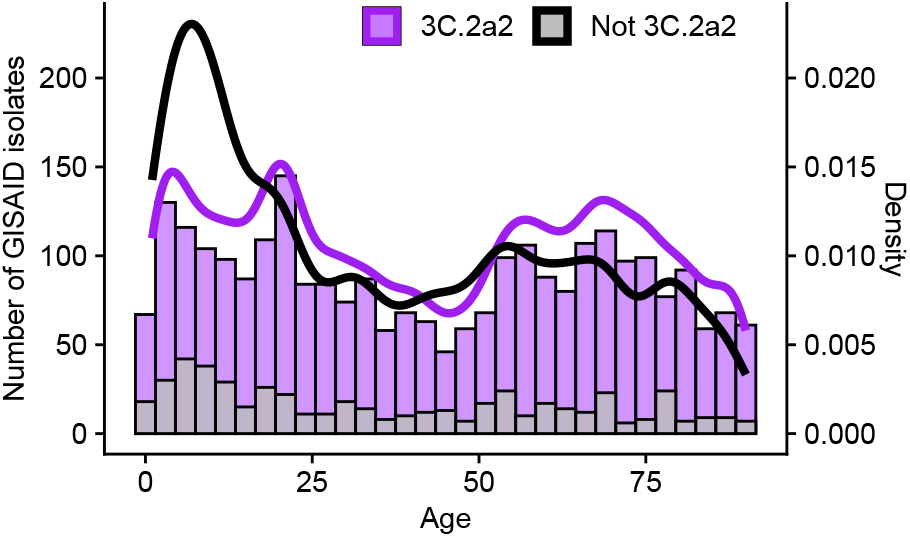
Host age distribution of H3N2 isolates sampled in the United States during the 2017/18 season. We obtained isolate data from GISAID. Viruses from clade 3C.2a2 are shown in purple, and viruses from all other clades combined are shown in gray (bars are overplotted). 3C.2a2 isolates were more common across all ages than viruses from other clades combined, but viruses from other clades were proportionally more common in children relative to adults.

### Correlations between titers to different strains vary by age, suggesting age-associated differences in epitope targeting

We next investigated the correlations in titers to different clades (Fig 5A): Do individuals with high titers to 3C.3a tend also to have high titers to 3C.2a2, for instance? Closely correlated titers to related viruses suggest that individuals might target epitopes conserved among them, which could underlie differences in neutralizing titers between age groups. (High titers to multiple strains could also indicate recent infections or immunizations with each of those strains and responses to their *non*-shared epitopes, although H3N2 infections typically occur at least several years apart and are less frequent in adults compared to children [17, 23, 24].) Aside from providing insight into the specificity of the antibody response, understanding the structure of titers within the population might lead to improved estimates of selective pressures on viruses. For instance, weakly correlated titers to different clades suggest a population with more heterogeneous immunity, which can affect viral coexistence, vaccination thresholds, and other dynamics [25–28].

**Fig 5.**
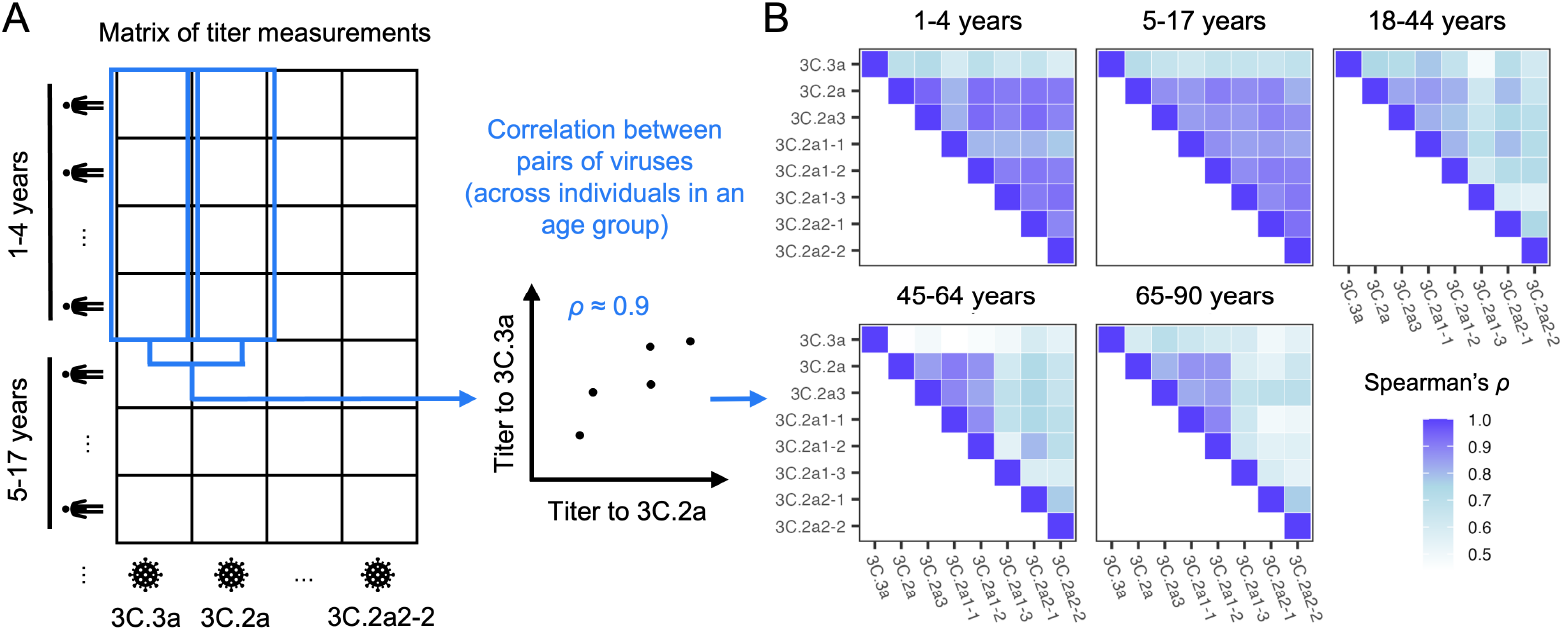
Correlations in titers to different clades. A. Schematics demonstrating how we calculated the correlation in titers to each strain pair across people in each age group. For these analyses, we randomly imputed continuous titer values between consecutive dilutions (e.g., a titer of 160 was replaced by a continuous value between 160 and 320 drawn with uniform probability). For each pair of viruses and each age group, we report the average Spearman correlation coefficient across 1000 replicate imputations. We removed individuals with undetectable titers across all reference viruses. B. Correlations between titers to different strains differ by age group, suggesting age-dependent patterns of epitope targeting.

After removing individuals with undetectable titers to all strains from the analysis, we found that the strength of correlation differed by age group and virus pair. In general, titers to all the reference viruses but 3C.3a were highly correlated in children and less correlated in older ages (Figs 5B, S16). This suggests that children target epitopes common to many reference viruses or have been infected by close relatives of each, whereas older age groups target epitopes conserved among only a subset. Results hold when age groups are chosen to span an equal number of years (Fig S17), showing that the weaker correlations in adults 18-44 y, 45-64 y, and 65-90 y are not due simply to the groups’ relative sizes or the diversity of childhood exposures represented in them. In contrast to children and younger adults, 45-to 90-year olds have their highest titers and strongest titer correlations to a distinct subset of reference strains (3C.2a, 3C.2a3, 3C.2a1-1, and 3C.2a1-2; Figs 2 and 5) that share many epitope A residues not conserved in the other strains (e.g. 128T, 131T, 135T, 138A, 142R), suggesting antibody responses focused on this antigenic region. In all age groups, titers to 3C.3a were least correlated with titers to other viruses (Fig S18). This might be explained by reduced exposure to 3C.3a viruses, especially in adults (Fig S3), and/or the targeting of sites on 3C.2a clade viruses that are not shared with 3C.3a (e.g., sites 128, 138, 142, and 144 in epitope A, the last potentially masked by a glycan in 3C.3a).

## Discussion

Current approaches for forecasting influenza and mapping its antigenic evolution rely on antigenic distance measurements that do not always reflect immunity in the human population. Understanding the size of the difference and how much it matters would require analyzing discrepancies between antibody titers and traditional ferret-based measurements over multiple years from representative cross-sectional surveys in different populations. Multiple years of sampling could also resolve the subpopulations and measures needed to assess immune selective pressures and compare them to other factors influencing fitness and growth rates [3, 29–33]. Here, as a proof of principle, we demonstrate how human sera can reveal differences in expected susceptibility to circulating HA clades that predict the clade circulating in the following season. The sera also demonstrate high heterogeneity in neutralizing titers by age. The consequences of these differences remain unclear, but they partly predict the relative susceptibility of different age groups to different clades in 2017-2018.

These findings might have been useful before the 2017-2018 influenza season in the United States. Although impractical to update the vaccine strain so near the start of the season, efforts to increase coverage to offset the expected low vaccine effectiveness might have blunted the season’s unusual severity, especially in the most vulnerable age groups. The 2017-2018 season caused approximately 41 million illnesses and 52,000 deaths [34]. The low effectiveness of the vaccine against H3N2 that season was attributed to egg adaptations that created a mismatch to circulating strains [35]. The H3N2 component of the vaccine, A/Hong Kong/4801/2014 (a basal 3C.2a strain), had been unchanged from the previous season because no clear indication of antigenic evolution was apparent by early 2017, when vaccine strain composition for the Northern Hemisphere was decided; the 3C.2a2 clade was nonetheless noted to be growing quickly [36]. Over 90% of 3C.2a2 strains isolated from the United States in the 2017-2018 season were described as well inhibited by ferret antisera raised against the cell-propagated reference virus for A/Hong Kong/4801/2014 (A/Michigan/15/2014), and in early 2018, the H3N2 vaccine component was updated only to avoid egg adaptations, not because antigenic change had been detected [37]. (Notably, a later investigation of H3N2 viruses circulating in Japan in 2017-2018 did detect antigenic differences between 3C.2a and 3C.2a2 strains using ferret antisera [38].) Our study shows that antigenic changes in fact were detectable in human sera by at least the summer of 2017, and they could predict the dominance of 3C.2a2 and the populations more susceptible to infection. Consistent with this prediction, Ursin et al. found that individuals testing positive for H3N2 in the 2017-2018 season had consistently lower serum neutralization titers to the 3C.2a2 clade than those testing negative—with no differences between the two groups’ titers to cell-grown A/Hong Kong/4801/2014—underscoring the consequences of these neutralization differences for protection and potentially transmission [39]. While our results suggest that the R261Q mutation contributed to 3C.2a2’s success, smaller clades that acquired the same mutation independently never rose to high frequencies.

Measurements of population immunity could be substantially more efficient and useful for forecasting if we understood exactly what to measure and in whom. Antibody titers to HA have been an established correlate of protection for half a century, and antibodies to NA for approximately a decade. The generally good concordance between hemagglutination inhibition assays and microneutralization suggest neutralization is a decent surrogate, but it is unclear how much protection each immune response confers in different people and whether measures of neutralization, total binding, antibody-dependent cellular cytotoxicity or phagocytosis activity, and/or potentially other B- or T-cell or innate immune measures could improve estimates of relative susceptibility. Correlates likely vary in quality over time and between age groups: large discrepancies between binding antibody titers and neutralization or protection have been reported and are associated with priming to other strains [17, 19, 40, 41]. Furthermore, immune measures associated with reduced transmission rather than simply protection against disease would provide more accurate estimates of viral fitness. It might also be important to weight immunity in different subpopulations differently: for instance, an infected child might be more likely to transmit than an infected adult. These considerations would affect the need to sample particular populations, such as unvaccinated members of certain age groups. Over larger geographic scales, samples from typical “source” populations may be better predictors or provide a longer lead time than populations that export fewer strains [42–44].

Evolutionary forecasts, including for vaccine strain selection, might benefit from such improved measures of population immunity but would require additional modifications. Clonal interference with multiple mutations can prevent any one clade from dominating a particular season [45–48] and complicate the relationship between susceptibility and clade frequency. Detailed clade frequency predictions might require incorporating estimates of population immunity into formal, potentially spatially explicit models that also consider other consequences of mutations on fitness [3, 49–51]. For influenza, estimates of viral fitness based purely on sequence data [49, 52] or on ferret-derived antigenic distances [3, 53] have modest success in out-of-sample forecasts, and models based on human population immunity have shown promise in predicting the frequencies of SARS-CoV-2 variants [50, 54]. Using clade frequencies to predict absolute disease burden would be a further challenge [55, 56]. To inform vaccine strain selection, human sera would need to be assayed much earlier relative to the influenza season than we did here. Perhaps most important, given variation in the immunogenicity and cross-immunity of different strains, it might not be optimal to be vaccinated against the strain forecasted to dominate [57].

Our data revealed variation in antibody titers between age groups that are broadly consistent with influenza’s epidemiology but lack precise explanation. Children over five years old had the highest geometric mean titers to all strains. This is consistent with the high attack rates in school-age children [58, 59] and other studies that report young children having the high titers to recent strains [60]. Children also had relatively high vaccination coverage (approximately 59% in the 2016-2017 season in children ≥ 6 mos.) compared to younger adults [61]. These two factors might interact, since recent infection can boost vaccine immunogenicity [62, 63]. The relatively high vaccination coverage in the oldest age group (approximately 65% in adults ≥ 65 y) might explain their higher titers compared to middle-aged and younger adults. Surprisingly, most middle-aged individuals had no detectable neutralizing antibodies to the HAs of circulating H3N2 clades. These results suggest antibodies to HA may be a poor correlate of protection in this age group and complement other reports of their discrepant anti-HA titers [19, 64]. We also observed that unlike children, adults had highly correlated titers to a subset of 3C.2a strains suggestive of antibody responses focused on epitope A. Consistent with this observation, Welsh et al. [33] recently applied deep mutational scanning to H3N2 and sera isolated in 2020. They found that compared to children, adults derived a larger fraction of their neutralizing response to epitope A, which had been immunodominant during the early 1990s [65].

Although not presented here, we fitted dozens of generalized linear mixed models to attempt to explain individuals’ titers to these strains as a function of potential recent infections, vaccinations, early infections with strains with homologous epitopes, and individual-specific biases in the contributions of different epitopes to titers. These models were inconclusive, suggesting a need for more careful study of how a person’s antibody titers change over time in response to exposures, and potentially with some deconvolution of the response to specific epitopes [66].

Our results demonstrate the feasibility of detecting significant differences in neutralizing titers to different H3N2 clades in a convenience sample of few hundred sera. This approach could entail substantial improvements over the use of ferret sera, which do not capture the immune history and heterogeneity in the human population [33]. Testing improved sampling protocols and forecasting models, which would be facilitated by the existence of global blood banks [67, 68] and common standards [69], might yield rapid advances in forecasting not only the dominant clade but also potentially the dominant subtype, and ideally at longer lead times than shown here. If linked to other forms of surveillance, cross-sectional sera might also help predict season severity and attack rates by age, as suggested here. The same samples and approximations of fitness might also predict the dynamics of other pathogens.

## Materials and methods

### Serological data

Sera from 489 individuals were collected between May and August of 2017 from the Children’s Hospital of Philadelphia (1-to 17-year-olds) and from the University of Pennsylvania Health care system via Penn BioBank (18-to 90-y-olds), as reported in [19]. Serum samples from children were leftover samples originally collected for lead testing and were de-identified for use in this study. We do not have access to clinical characteristics of the donors. The Penn BioBank routinely collects serum samples from individuals visiting the University of Pennsylvania Health care system. We did not include samples collected by the Penn BioBank from donors who had a pregnancy reported during the last 9 months, who had a medical history of cancer or organ transplantation, or who had a reported infectious disease within the previous 28 days. The study complied with all relevant ethical regulations and was approved by the Institutional Review Board of the University of Pennsylvania. Leftover de-identified samples collected at CHOP were considered exempt from human research (exemption 4) since the samples were leftover discarded samples that were completely de-identified before our research team received them.

We performed foci reduction neutralization tests (FRNT) on 437 individuals’ sera using the 8 HA reference viruses (3C.3a, 3C.2a, 3C.2a1-1, 3C.2a1-2, 3C.2a1-3, 3C.2a2-1, 3C.2a2-2 and 3C.2a3), and enzyme-linked lectin assays (ELLA) on 352 individuals using the two NA reference viruses (3C2.A (NA) and 3C.2a2-2 (NA)) as described in [19]. Full experimental details can be found in [19]. Briefly, for FRNT, we treated serum samples with receptor-destroying enzyme, serially diluted them twofold, incubated them with virus at a concentration of ≈ 300 focus-forming units for 1 h at room temperature, and added the mixture to confluent MDCK-SIAT1 cell monolayers in a 96-well plate. We incubated the cells with the virus-serum mixture in 5% CO_2_ for 1h at 37 ºC, then washed them with Minimal Essential Media (MEM). After additional steps [19], we imaged the plates to quantify foci using an ELISpot reader. We report FRNT titers as the reciprocal of the highest dilution of sera that reduced the number of foci by at least 90% relative to control wells with no serum. We assigned a titer of 10 to serum samples that failed to achieve a 90% reduction at the smallest dilution (1:20). For ELLA, we incubated virus and diluted heat-inactivated serum overnight at 37 ºC in microtiter 96-well plates coated with fetuin diluted in coating solution, washed them and performed additional steps before reading the plates at an OD of 450 nm using a microplate reader. We report ELLA titers as the reciprocal of the highest dilution of sera that reduced the OD value by at least 50%, relative to control wells with no serum and after background subtraction. We assigned a titer of 10 to serum samples that did not show at least 50% OD reduction at the smallest dilution (1:20). There were no significant titer differences between batches.

We visualized titers to different reference strains by age using locally estimated scatterplot smoothing (LOESS) curves with a smoothing parameter *α* = 0.75 and degree = 2.

### Viruses

We generated all viruses using reverse genetics, as described in [19]. Briefly, we cloned HA and NA genes for the reference strains into the pHW2000 reverse-genetics plasmid along with internal genes from A/Puerto Rico/8/1934. For viruses used in the FRNT assay, we used the NA gene of A/Colorado/15/2014. For viruses used in ELLA, we used an H6 gene from A/turkey/Massachusetts/3740/1965 and the NA gene from either A/Colorado/15/2014 (representing clade 3C.2a) or A/Pennsylvania/49/2018 (representing 3C.2a2-2). We transfected the plasmids into co-cultures of 293T and Madin-Darby Canine Kidney (MDCK)-SIA1 cells, harvesting supernatants 3 days after transfection and storing them at -80 ºC. We sequenced HA and NA genes to confirm that no additional mutations arose during transfection.

### Genealogy of H3N2 and clade-specific amino acid substitutions

Prior to our analyses, we downloaded all available H3N2 HA and NA sequences from the 2012-13 season through the 2017-18 season from GISAID (accessed in 01/10/2022). Sequences were aligned using MAFFT 7.310 [70].

We downsampled sequences to construct the phylogeny. From the 2012-13 through the 2015-16 season, we sampled 20 sequences per season. For the 2016-17 and 2017-18 season, 100 sequences were sampled per season. The GISAID accession IDs and metadata of the sequences used for the analysis are available in the Supporting Information. We used BEAST 2.6.6 to reconstruct the genealogy [71] with a HKY substitution model [72] with a four-category gamma site model with 4 and a log normal relaxed clock. A coalescent Bayesian Skyline tree was used for the prior. We ran the chain for 50 million steps and saved every 1000 trees, using 5 million steps as burn-in. The maximum clade credibility tree was obtained using TreeAnnotator 2.6.6 version.

To visualize the tree, we used the R package ggtree 3.0.4 [73]. The trees were colored by clade. For the genealogy of the 2016-17 season, only tips of sequences collected in North America during the 2016-17 season were shown; these circled tips are colored according to their assigned clade. For sequences collected in other areas or seasons, only branches were shown. Similarly, for the genealogy of the 2017-18 season, only sequences collected in North America in that season are shown as colored circles.

Sequence samples were assigned to reference viruses according to reference virus-specific mutations at segregating sites, shown in Fig 1B. Here, sequences were assigned to each reference virus rather than the subclade represented by each reference strain. This is because sequences with 171K, 121K, and 135K, such as reference strain 3C.2a1-3, occur multiple times in clade 3C.2a1, and thus these sequences do not belong to any one subclade of 3C.2a1. Additionally, within a subclade, mutations at segregating sites occur so that a sequence in the same clade as a reference virus may not share the same genetic characteristics. Due to frequent mutation at residue 142 across most of clades, we allowed residue 142 to have any amino acid across most of clades, except for clade 3C.2a2, which has a clade-specific 142K substitution. We confirmed that all the sequences assigned to a reference virus fall in the same subclade as the reference virus.

Clade-specific substitutions were colored by epitope on the H3 structure using PyMOL version 2.3.3 [74].

### Inferring relative susceptibility from titers

We defined the relative susceptibility to a strain as the fraction of the population with titers to that strain below some threshold (here, initially 40 for HA and 80 for NA). To estimate this fraction for the U.S. population, we first computed the fraction below the threshold in different age bins in our sample. We then computed a weighted sum in which the weights were the projected fractions of the U.S. population in each age bin in 2017. This adjustment was necessary to obtain a representative estimate of immunity in the overall population, since sample availability varied by age. Suppose *S*_*i*_ is the fraction of the overall U.S. population susceptible to strain *i* and *Ŝ*_*i*,*a*_ is the fraction of serum samples in age bin *a* with titers to *i* below the threshold. Then

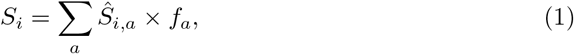

where *f*_*a*_ is the projected fraction of the U.S. population in age bin *a* in 2017. We started with age bins at the resolution of one year using data from [75]. When there were fewer than eight titer measurements for a year of age, that age was grouped with the next year of age to form a larger bin, and so on until the bin contained at least eight titer measurements.

To estimate the susceptible fraction for a particular age group in the U.S. population, we simply computed the sum above across age bins contained in that larger group, divided by the fraction of the U.S. population in those bins combined. For instance, to calculate the susceptible fraction among 5-17 year-olds in the U.S., we used

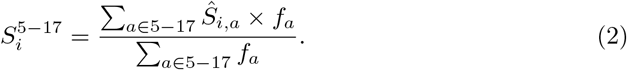

We found that using alternate titer thresholds for HA (Figs S6, S7, and S8) and NA (Figs S12 and S13) resulted in consistent relative susceptibilities across strains.

We alternatively measured relative susceptibility by the geometric mean titer (GMT). The GMT was weighted analogously by the population fraction of each age bin. Because lower GMTs correspond to higher susceptibility, we used a reverse scale when showing the relative susceptibility by GMT.

To test for meaningful differences in relative susceptibilities, we bootstrapped individuals to determine if the difference in inferred relative susceptibilities between two viruses was significantly greater than zero [76]. For each age bin, individuals were resampled 1000 times with replacement, and the fraction of individuals susceptible to each virus was calculated. For a given pair of viruses, we defined the relative susceptibility difference observed in the data as 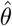. The bootstrap value of 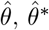, was obtained 1000 times by resampling individuals. Then we obtained the null distribution of 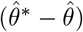 and calculated the probability (*p*) of observing 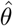 or a greater value under this null distribution. If *p <* 0.05 (with no correction for multiple testing), the relative susceptibility difference is significantly greater than zero, i.e., susceptibility to the first virus significantly exceeded that to the second virus. For a given virus, we perform this comparison against all other viruses and counted the number of significant results. The more significant results, the lower the rank (closer to 1) of the relative susceptibility to a virus. We used the same approach and significance level for all other bootstrapping analyses.

### Frequencies of subclades

To calculate the frequencies of different subclades, we downloaded sequences from the 2016-17 to the 2018-19 seasons available on GISAID on January 10, 2022, and assigned sequences to each subclade using the same method as was used to construct the genealogy. Because there were few sequences from Philadelphia, we calculated subclade frequencies in three different ways, using sequences collected from North America, United States, or the northeastern US. We considered Region 1, Region 2, and Region 3 of the U.S. Outpatient Influenza-like Illness Surveillance Network (ILINet, [61]) as the Northeastern U.S. states. These states are Connecticut, Maine, Massachusetts, New Hampshire, Rhode Island, Vermont, New Jersey, New York, Delaware, the District of Columbia, Maryland, Pennsylvania, Virginia, and West Virginia. Region 2 of ILINet includes Puerto Rico and the Virgin Islands, but we excluded them from the analysis of the northeastern U.S. For estimates derived from North American sequences, we used 4488 and 5425 sequences from the 2016-17 and 2017-18 seasons, respectively. For the US, 3707 and 3782 sequences were used. For the northeastern US, 782 and 676 sequences were used. The GISAID accession IDs and metadata of the sequences used for the analysis are available in the Supporting Information.

### Correlations between titers to different strains

For each age group and pair of viruses, we calculated Spearman’s *ρ* using the cor function in R. To account for the interval censoring of titer data and the presence of a lower limit of detection, we randomly imputed continuous titer values and calculated the average regression coefficient across 1000 imputations. Titers below the lower limit of detection (1:20) were uniformly sampled between the lowest possible titer (1:1, indicating no dilution) and 1:20. Titers at or above the limit of detection were randomly sampled from the interval between the recorded titer and the next dilution (e.g., a recorded titer of 1:160 was imputed a value between 1:160 and 1:320, with uniform probability). For this analysis, we excluded individuals with undetectable titers to all strains. We excluded 45/112 (40%) of *<*5 y olds, 10/164 (6%) of 5-17 y olds, 31/89 (35%) of 18-44 y olds, 28/62 (45%) of 45-64 y olds, and 22/62 (36%) of 65-90 y olds. For each virus pair, we tested the difference in correlation coefficients between the youngest age group and each other age group using the same bootstrapping procedure we used to test for differences in susceptibility among strains within an age group (Fig S16).

We also used bootstrapping to evaluate differences in correlation coefficients between viral pairs within an age group. For each virus pair, we did a series of bootstrap tests comparing the pair’s correlation coefficient with the coefficient for each of the other pairs. Then, for each virus pair, the number of tests in which the pair’s correlation was significantly weaker than that of other pairs within the group was counted. In each age group, there are 28 virus pairs whose correlation coefficient was calculated. One of the pairs, for example, is 3C.3a and 3C.2a, and this pair’s correlation coefficient is compared with the other 27 correlation coefficients of other virus pairs. The 3C.3a v. 3C.2a pair’s correlation was weaker than 15 other pairs’ correlations. This number of tests in which the pair’s correlation was significantly weaker than other pairs within the group is shown as the color intensity of the heat map of Fig S18.

For these boostrapping procedures, we used a significance level of 95% without correction for multiple testing, randomly imputing continuous titers once for each bootstrap replicate.

## Data Availability

All data and code used in this analysis are available at https://github.com/cobeylab/population_immunity_predicting_flu.git. The sequences obtained from GISAID and used for the analyses can be accessed using the accession IDs provided in the Supporting Information.

## Acknowledgments

We thank Ed Baskerville for extensive help with analytic models that were ultimately not included in this paper but shaped the direction of the work. We thank anonymous reviewers, Manon Ragonnet-Cronin, Richard Neher, Jesse Bloom, and Tal Einav for comments on the manuscript. We gratefully acknowledge all GISAID data contributors, i.e., the authors and their originating laboratories responsible for obtaining the specimens, and their submitting laboratories for generating the genetic sequence and metadata and sharing via the GISAID Initiative, on which this research is based.

## Author contributions

Conceptualization–SC, SH. Formal analysis–KK, MCV, SC. Investigation–SG, MW. Supervision–SC, SH. Writing, original draft preparation–KK, MCV, SC. Writing, review and editing–KK, MCV, SG, SH, SC.

## Data and software availability

All data and code used in this analysis are available at https://github.com/cobeylab/population_immunity_predicting_flu.git. The Sequences obtained from GISAID and used for the analyses can be accessed using the accession IDs provided in Supporting information.

## Supporting information

**S1 File. GISAID acknowledgment table**

**S2 File GISAID accession IDs and metadata of the sequences used for constructing genealogy**

**S3 File GISAID accession IDs and metadata of the sequences used in the analysis of subclade frequency**

## Supplementary Tables and Figures

**Table S1.**
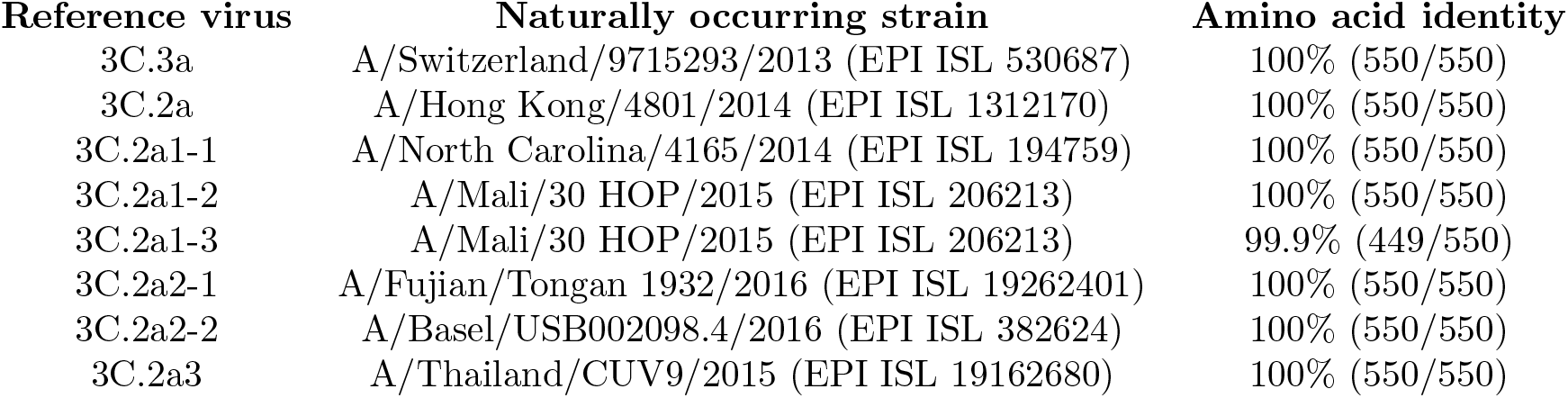
Naturally occurring strains with identical or nearly identical amino acid sequences to reference viruses.

**Fig S1.**
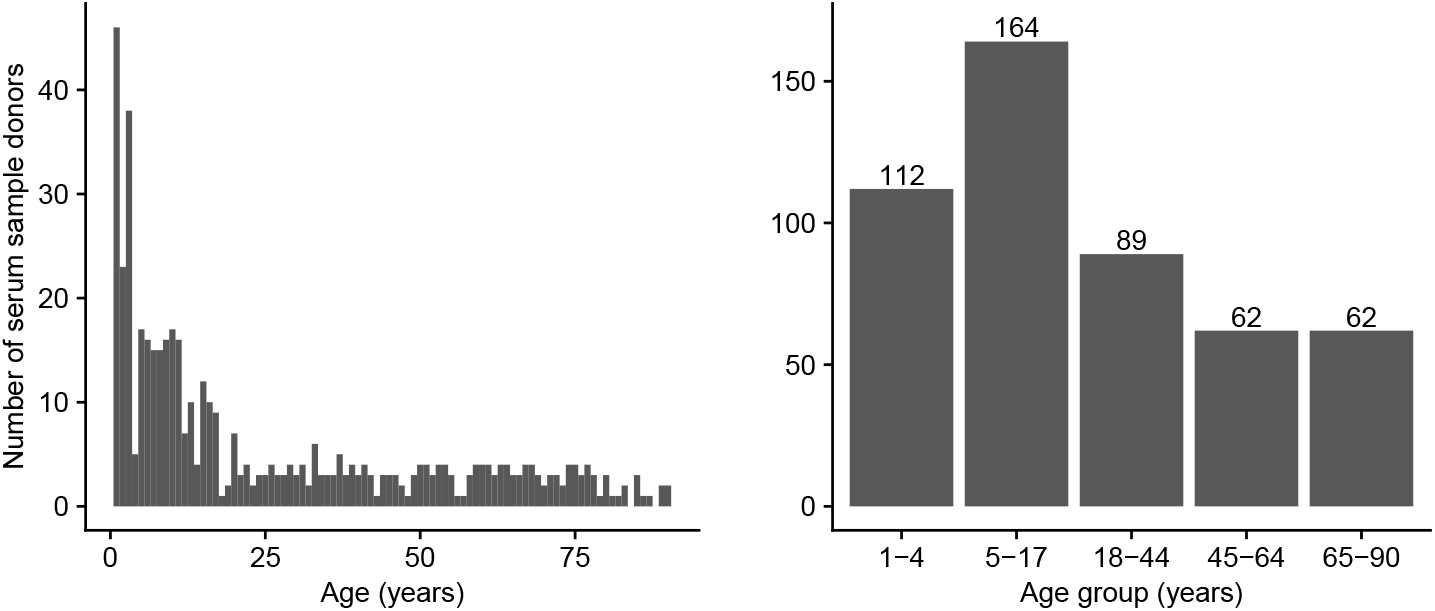
Age distribution of serum sample donors. We collected serum samples from May to August of 2017 from the University of Pennsylvania BioBank and Children’s Hospital of Philadelphia. The age distribution of donors at 1 year resolution is shown on the left, and the sizes of different age groups are shown on the right.

**Fig S2.**
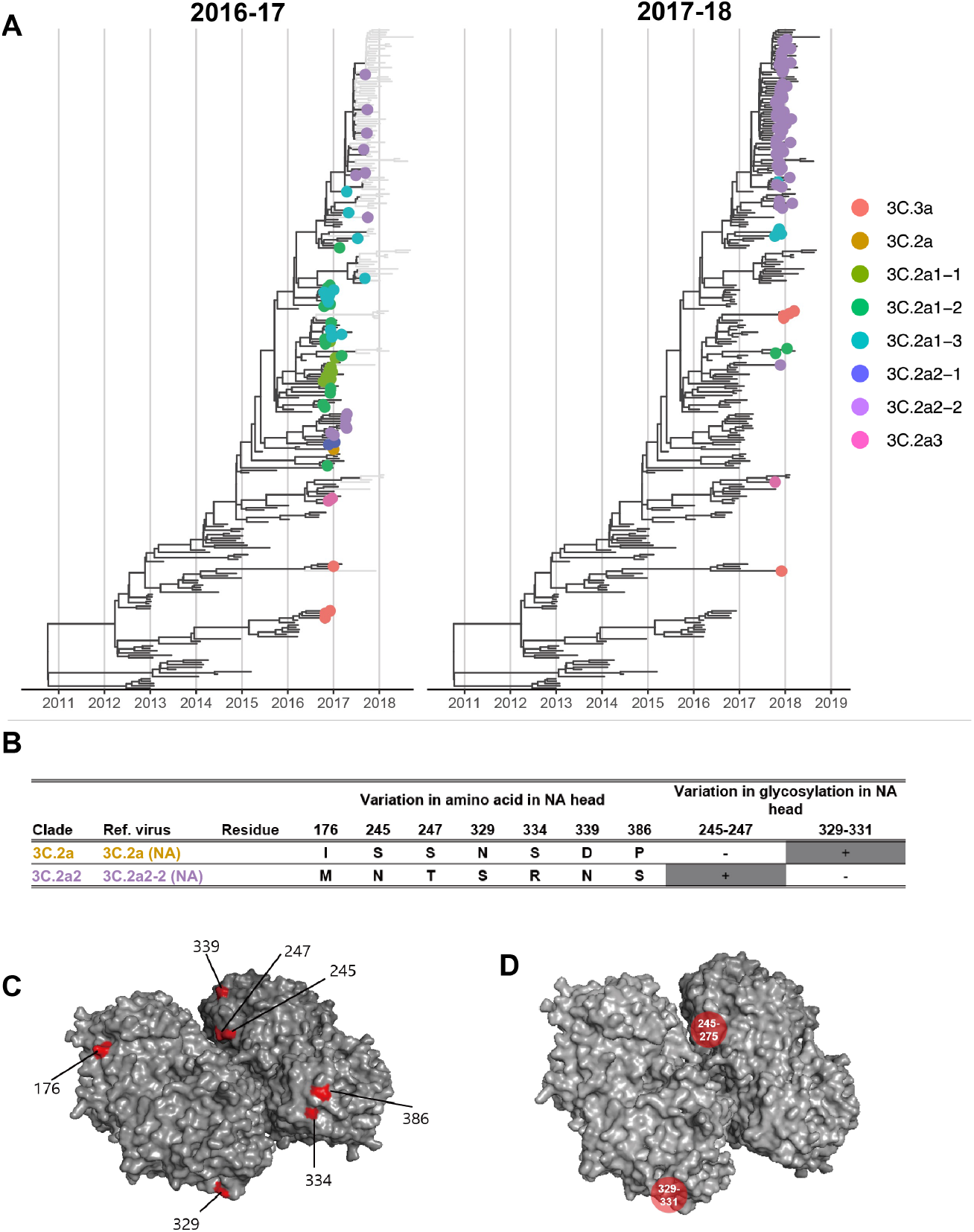
NA of co-circulating H3N2 clades during the 2016-17 and 2017-18 seasons. A. Genealogy of H3N2 showing NA sequence samples through the 2016-17 season (left) and through the 2017-18 season (right). Tips are filled circles if collected in North America during the 2016-17 season (left) and the 2017-18 season (right) and colored according to the associated test virus. B. Variable amino acids and PNGS between test viruses. Between clade 3C.2a (NA)and clade 3C.2a2 (NA), only substitutions at NA head are shown. C. The differences between clade 3C.2a (NA) and 3C.2a2-2 (NA) are shown on N2 head structure (Protein Data Bank: 6n4d). Amino acid differences between the two test viruses are colored in red. D. PNGS that vary between 3C.2a (NA) and 3C.2a2 (NA) are shown on the N2 structure.

**Fig S3.**
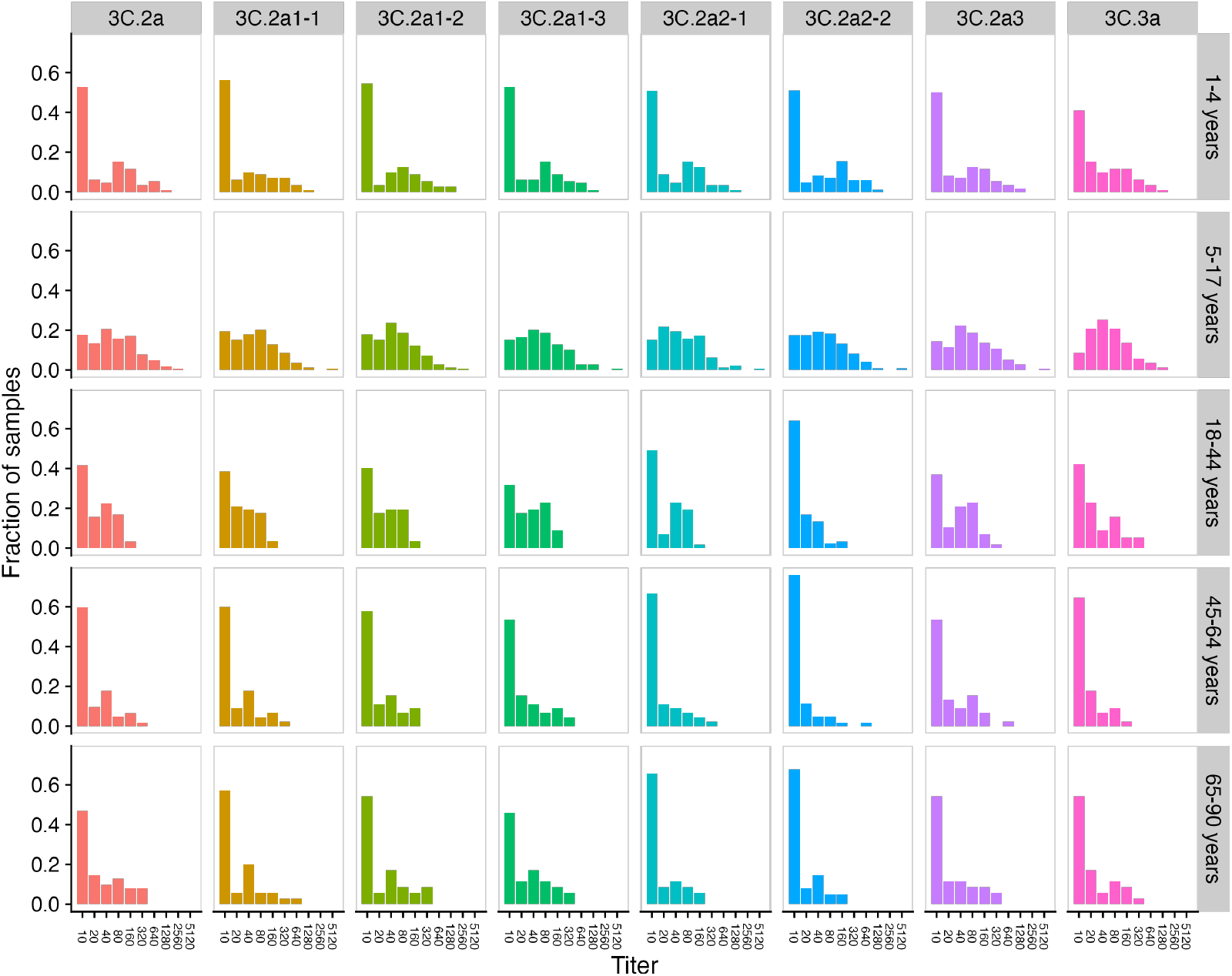
Distribution of HA titers for each strain by age group. A titer of 10 indicates the titer is below the limit of detection.

**Fig S4.**
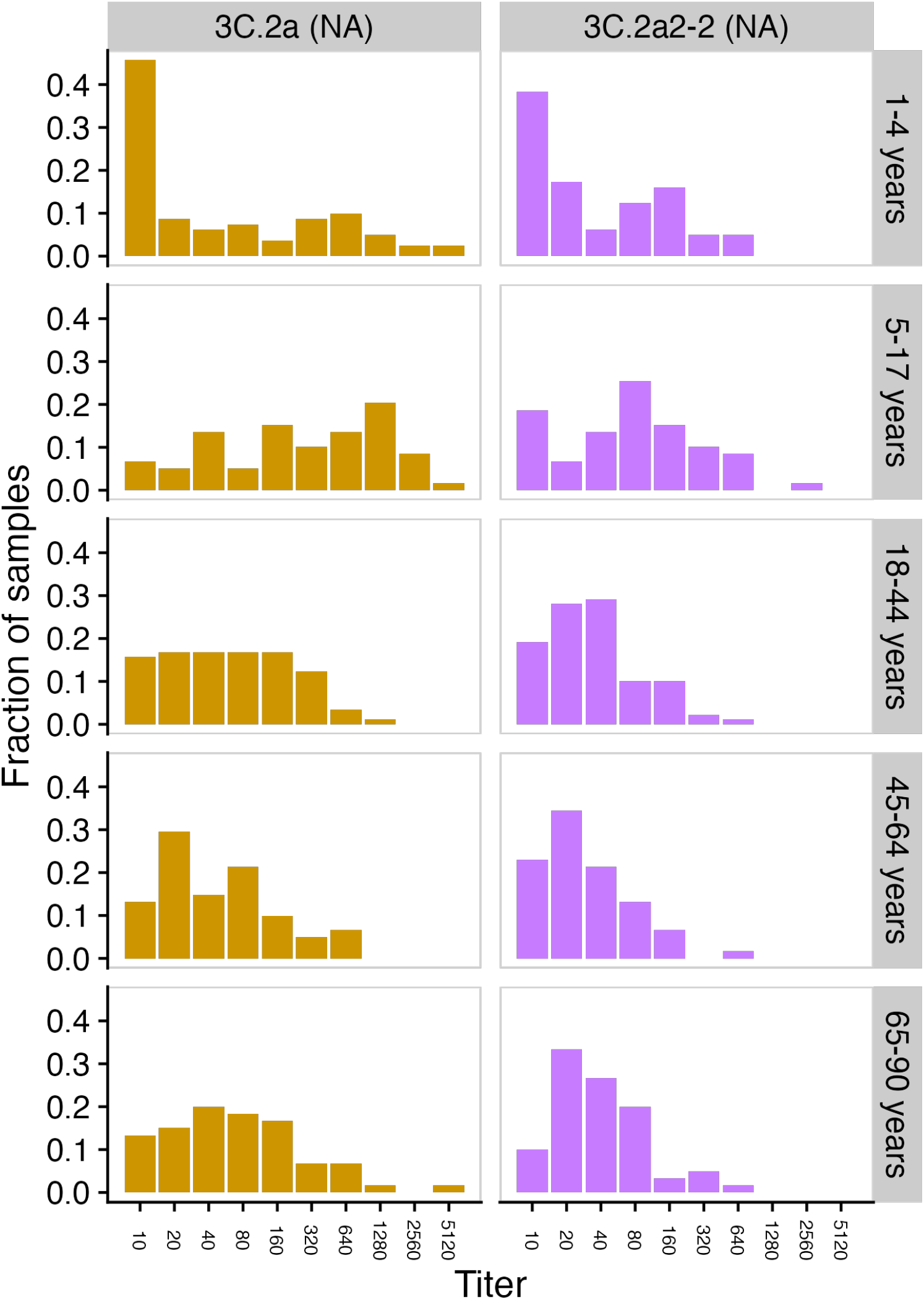
Distribution of NA titers for each strain by age group. A titer of 10 indicates the titer is below the limit of detection.

**Fig S5.**
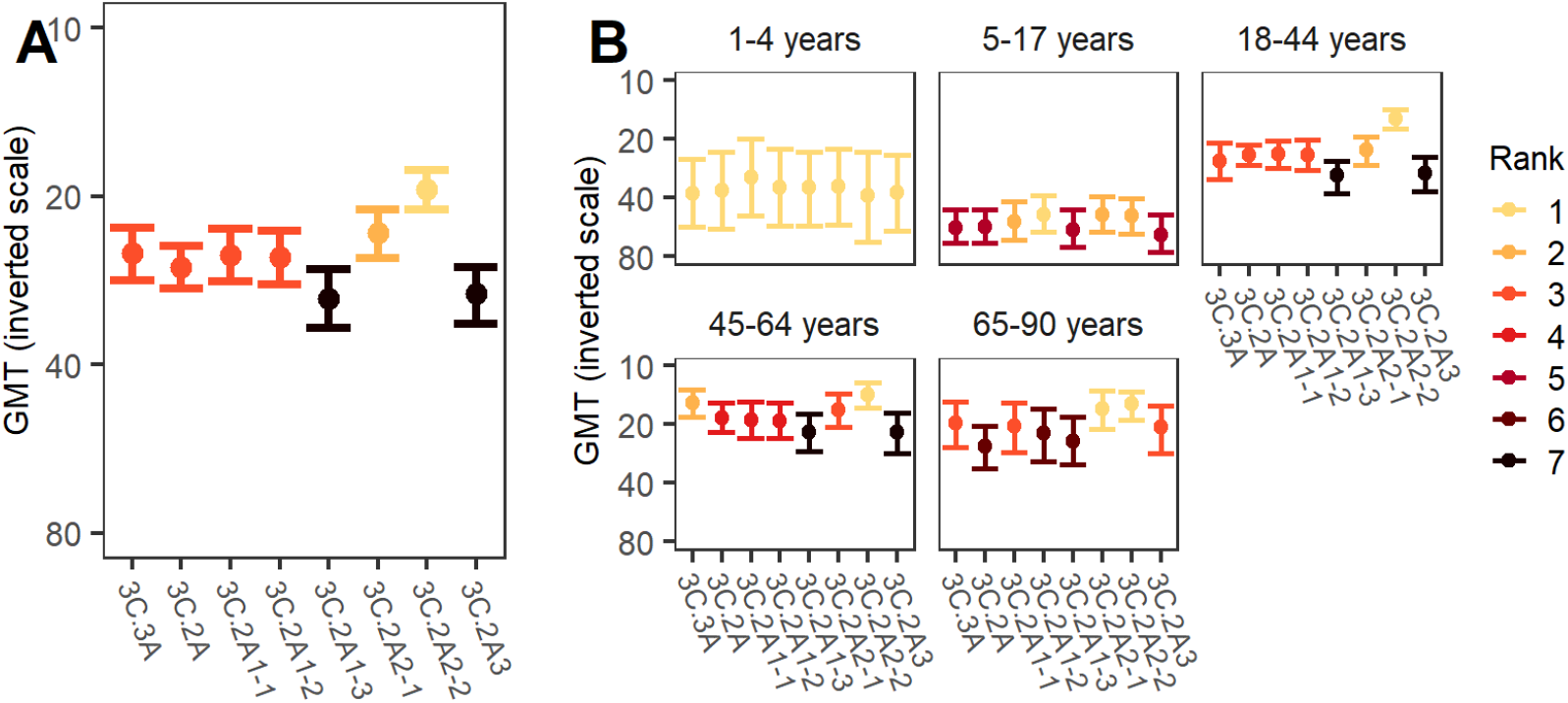
The GMT for each HA reference strain. Since lower GMT corresponds to higher susceptibility, we use an inverse scale to indicate the relative susceptibility. GMTs are shown for the whole population (A) and by age group (B). The bars indicate 95% CIs obtained from bootstrapping. A lower rank indicates significantly higher implied susceptibility: the strain with lowest GMT has rank 1, and strains are tied in rank and appear in the same color if their relative susceptibilities do not differ significantly.

**Fig S6.**
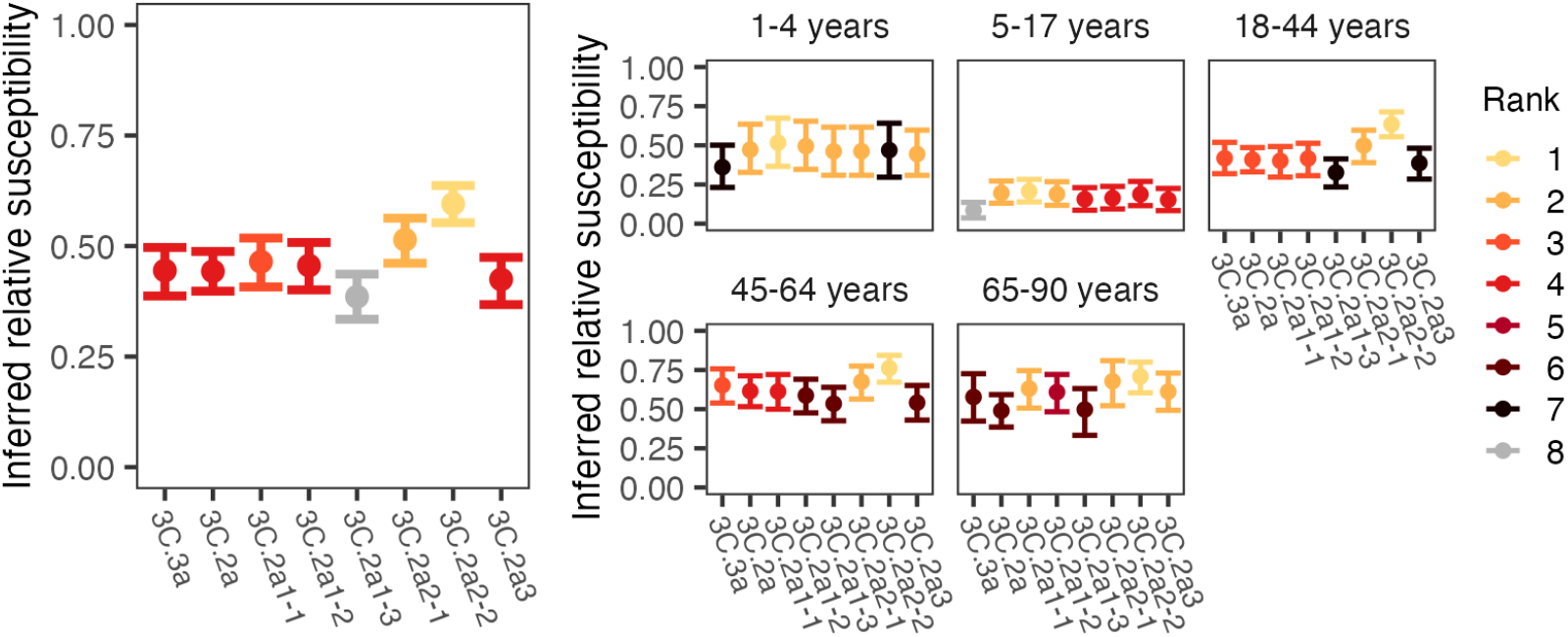
Inferred relative susceptibility to each reference strain using a 1:20 HA titer threshold. Inferred relative susceptibility and the susceptibility rank of each reference strain for the whole population (left) and by age group (right). The bars indicate 95% CIs obtained from bootstrapping. A lower rank indicates significantly higher susceptibility: the strain to which susceptibiity is highest has rank 1, and strains are tied in rank and appear in the same color if their relative susceptibilities do not differ significantly.

**Fig S7.**
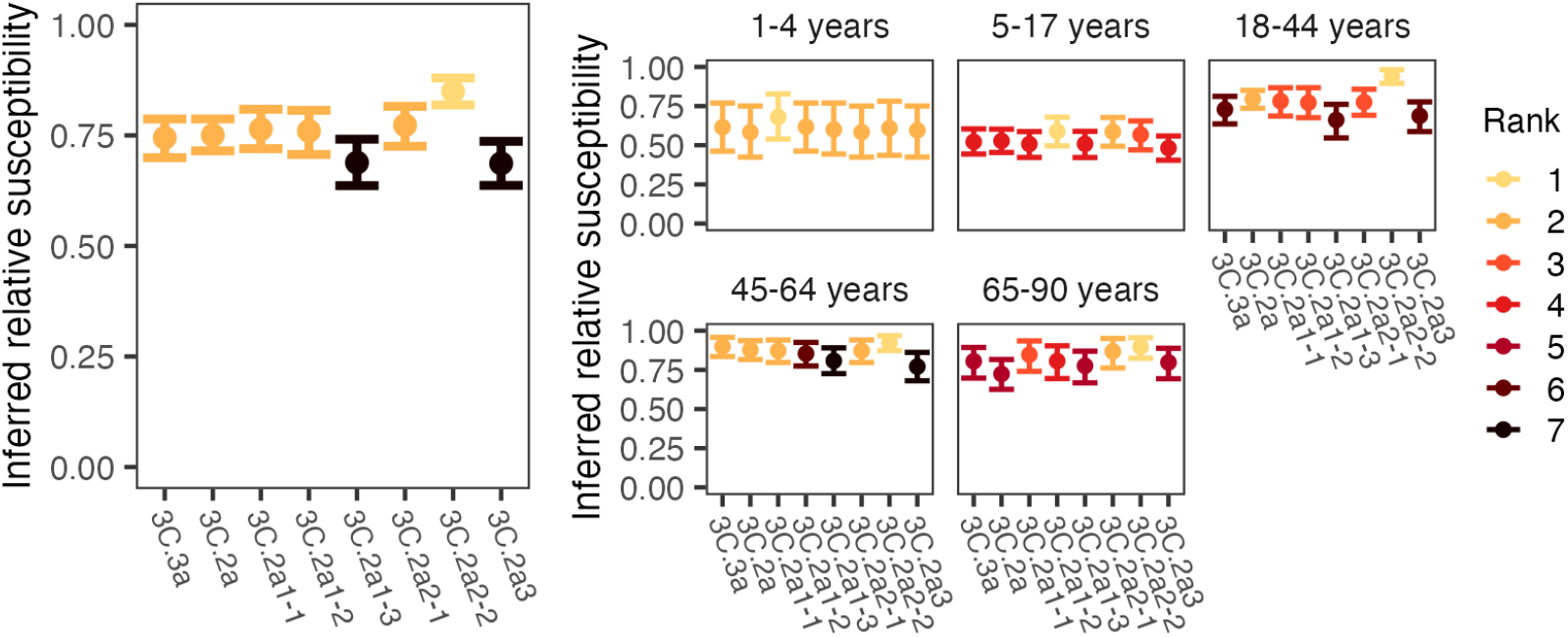
Inferred relative susceptibility to each reference strain using a 1:80 HA titer threshold. Inferred relative susceptibility and the susceptibility rank of each reference strain for the whole population (left) and by age group (right). The bars indicate 95% CIs obtained from bootstrapping. A lower rank indicates significantly higher susceptibility: the strain to which susceptibiity is highest has rank 1, and strains are tied in rank and appear in the same color if their relative susceptibilities do not differ significantly.

**Fig S8.**
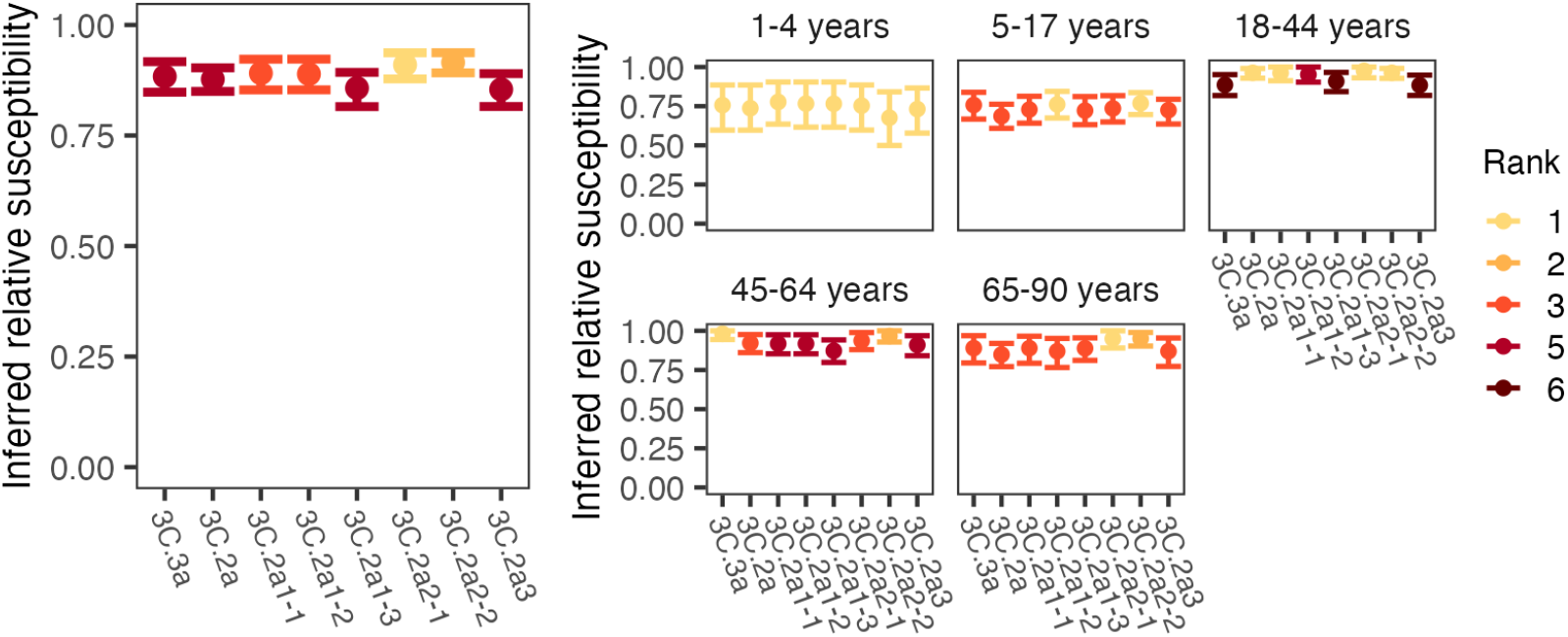
Inferred relative susceptibility to each reference strain using a 1:160 HA titer threshold. Inferred relative susceptibility and the susceptibility rank of each reference strain for the whole population (left) and by age group (right). The bars indicate 95% CIs obtained from bootstrapping. A lower rank indicates significantly higher susceptibility: the strain to which susceptibiity is highest has rank 1. Strains are tied in rank and appear in the same color if their relative susceptibilities do not differ significantly..

**Fig S9.**
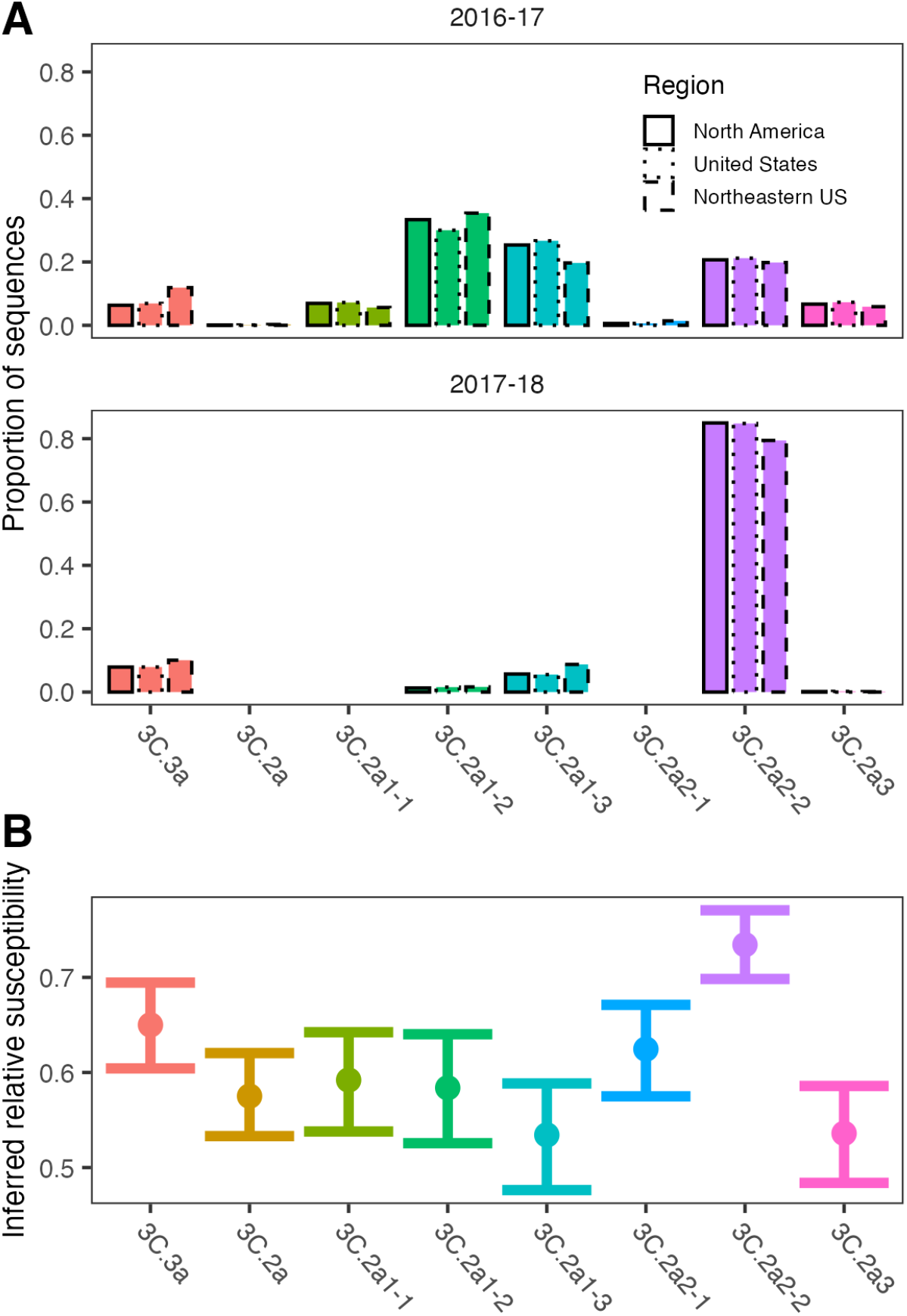
Comparison of H3 proportions by clade (reference strain) and inferred relative susceptibilities. A. Proportion of H3 sequences assigned to each test virus were calculated using GISAID sequences from North America, US, and the Northeastern US during the 2016-17 and 2017-18 season. B. Inferred relative susceptibilities to test viruses assuming a 1:40 HA titer threshold for protection.

**Fig S10.**
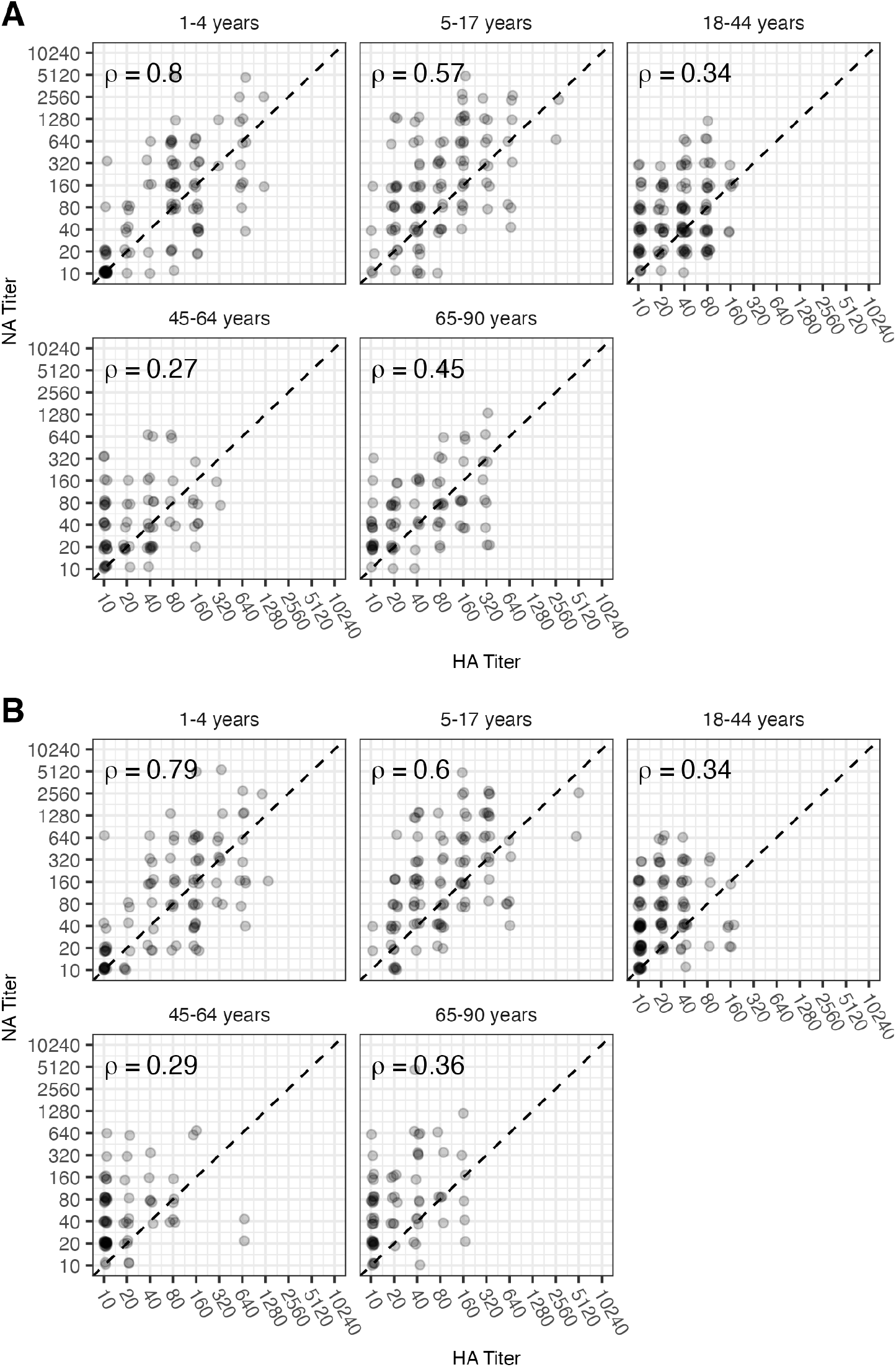
Correlations between HA and NA titers for 3C.2a (A) and 3C.2a2 (B). Points have been slightly jittered to show density. We computed the Spearman correlation coefficient separately for each strain and age group.

**Fig S11.**
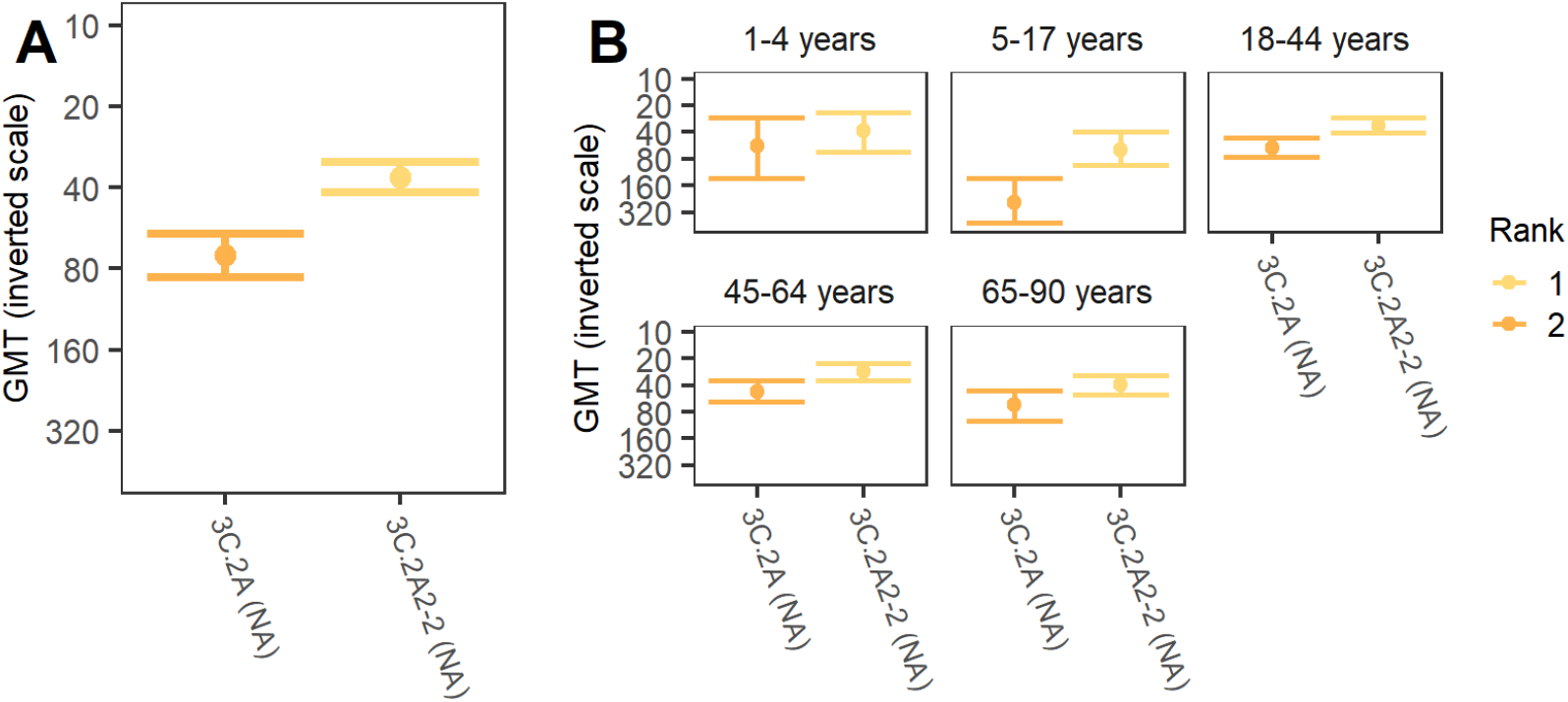
The GMT for each NA reference strain. Since lower GMT corresponds to higher susceptibility, we use an inverse scale to indicate the relative susceptibility. GMTs are shown for the whole population (A) and by age group (B). The bars indicate 95% CIs obtained from bootstrapping. A lower rank indicates significantly higher implied susceptibility: the strain with lower GMT has rank 1.

**Fig S12.**
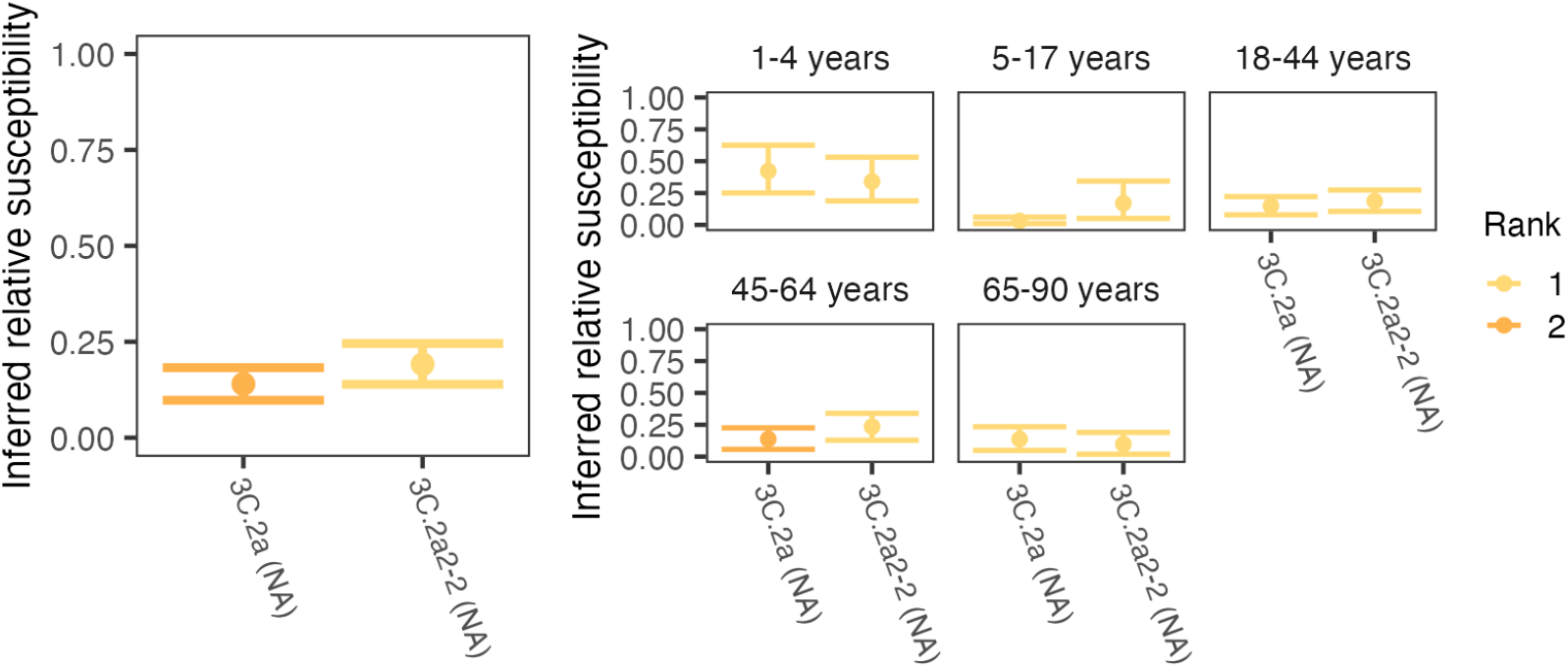
Inferred relative susceptibility to each NA reference strain using a 1:20 threshold. Inferred relative susceptibility and the susceptibility rank of each reference strain for the whole population (left) and by age group (right). The bars indicate 95% CIs obtained from bootstrapping. A lower rank indicates significantly higher susceptibility: the strain to which susceptibiity is highest has rank 1. Strains are tied in rank and appear in the same color if their relative susceptibilities do not differ significantly.

**Fig S13.**
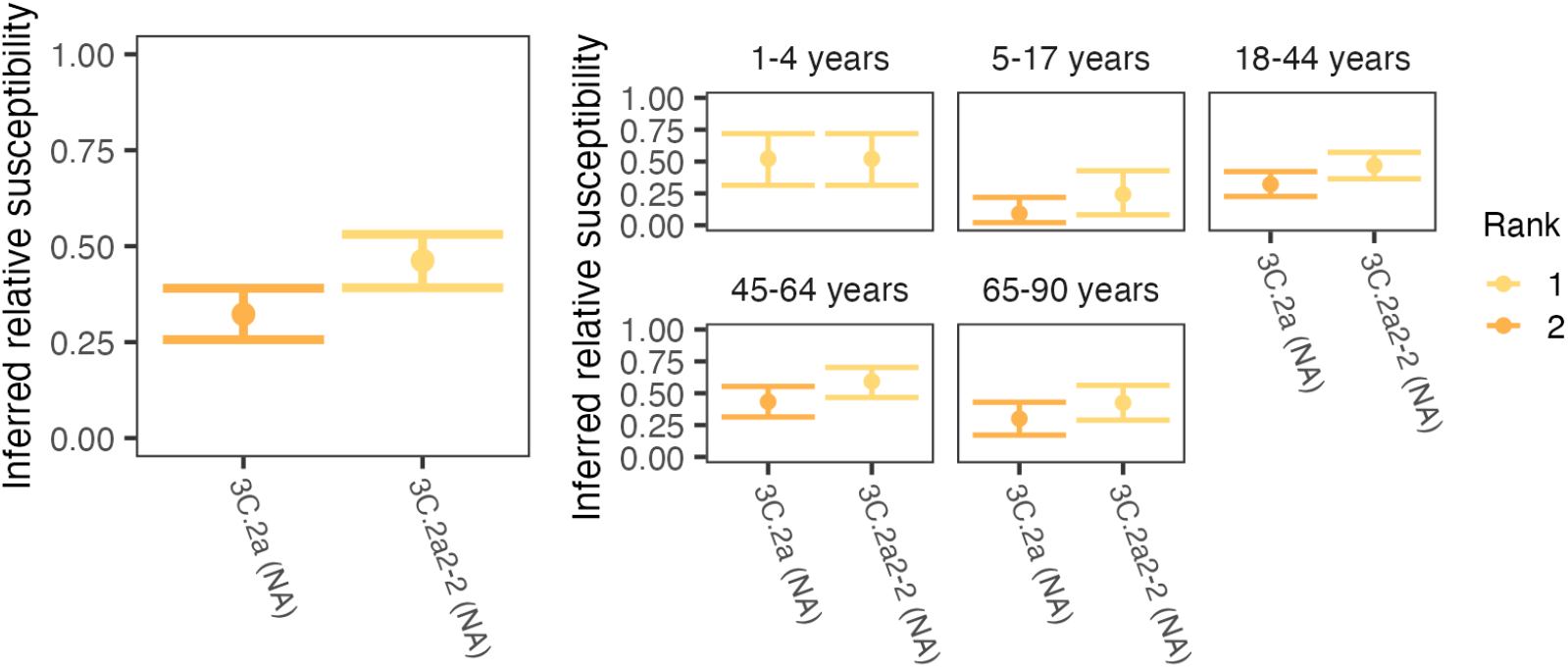
The inferred relative susceptibility to each reference strain using a 1:40 NA titer threshold. Inferred relative susceptibility and the susceptibility rank of each reference strain for the whole population (left) and by age group (right). The bars indicate 95% CIs obtained from bootstrapping. A lower rank indicates significantly higher susceptibility: the strain to which susceptibiity is highest has rank 1. Strains are tied in rank and appear in the same color if their relative susceptibilities do not differ significantly.

**Fig S14.**
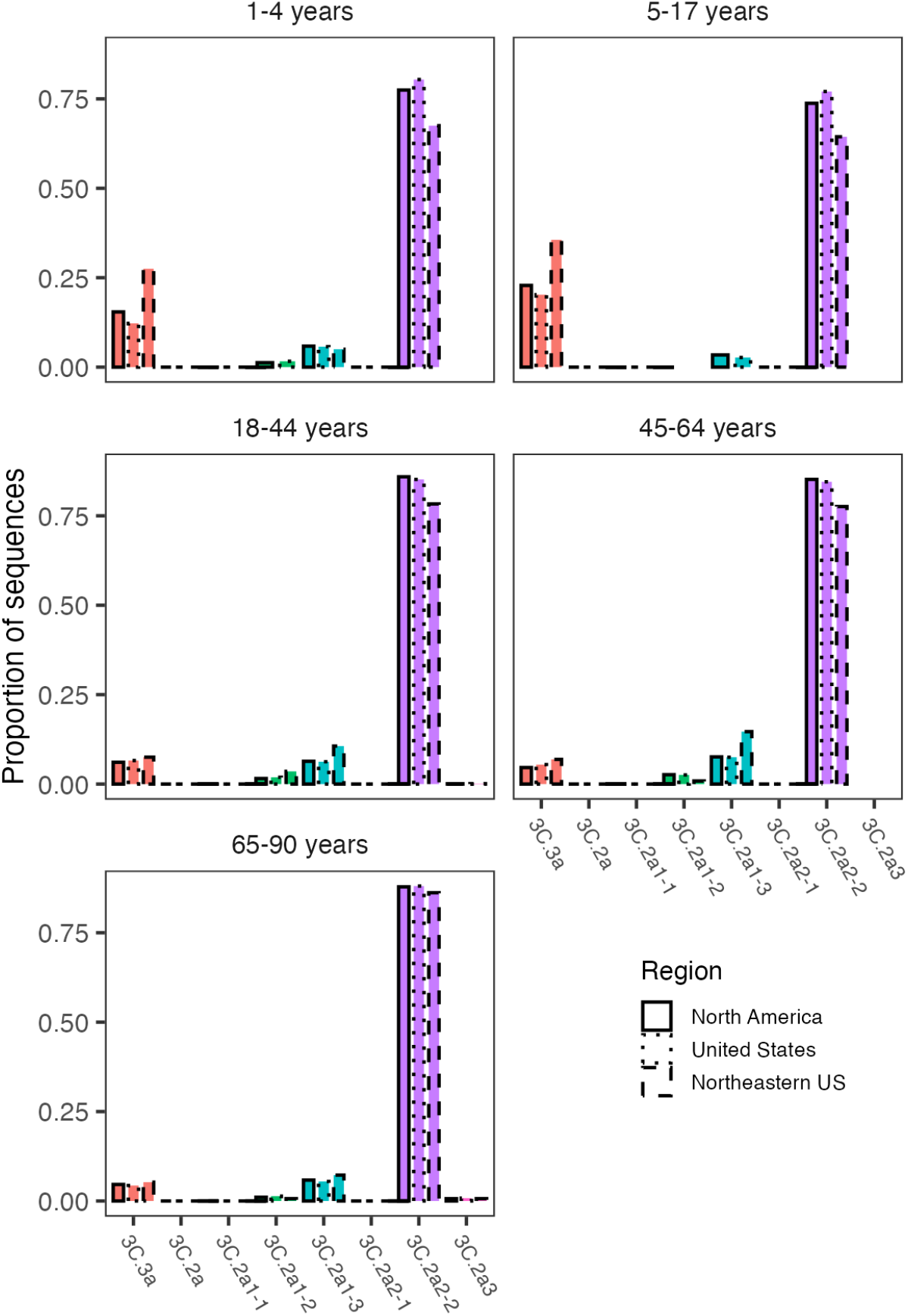
Proportions of H3 sequences in the 2017-18 season assigned to each reference virus by age group. Proportions were calculated using GISAID sequences collected in North America, United States, and the northeastern United States, respectively.

**Fig S15.**
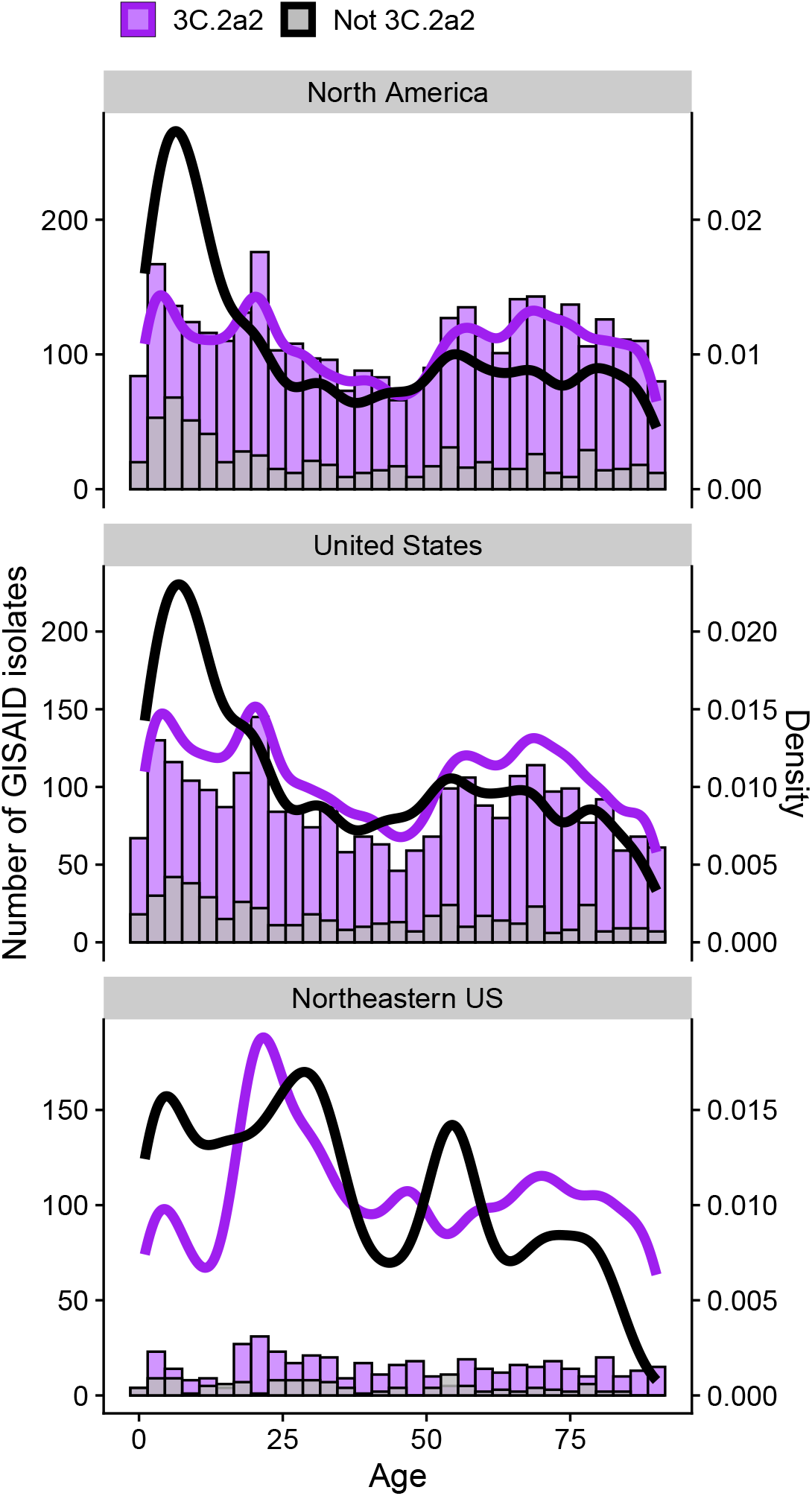
Host age distribution of H3N2 isolates sampled during the 2017/18 season, shown at different geographical resolutions. We obtained isolate data from GISAID. Viruses from clade 3C.2a2 are shown in purple, and viruses from all other clades combined are shown in gray (bars are overplotted).

**Fig S16.**
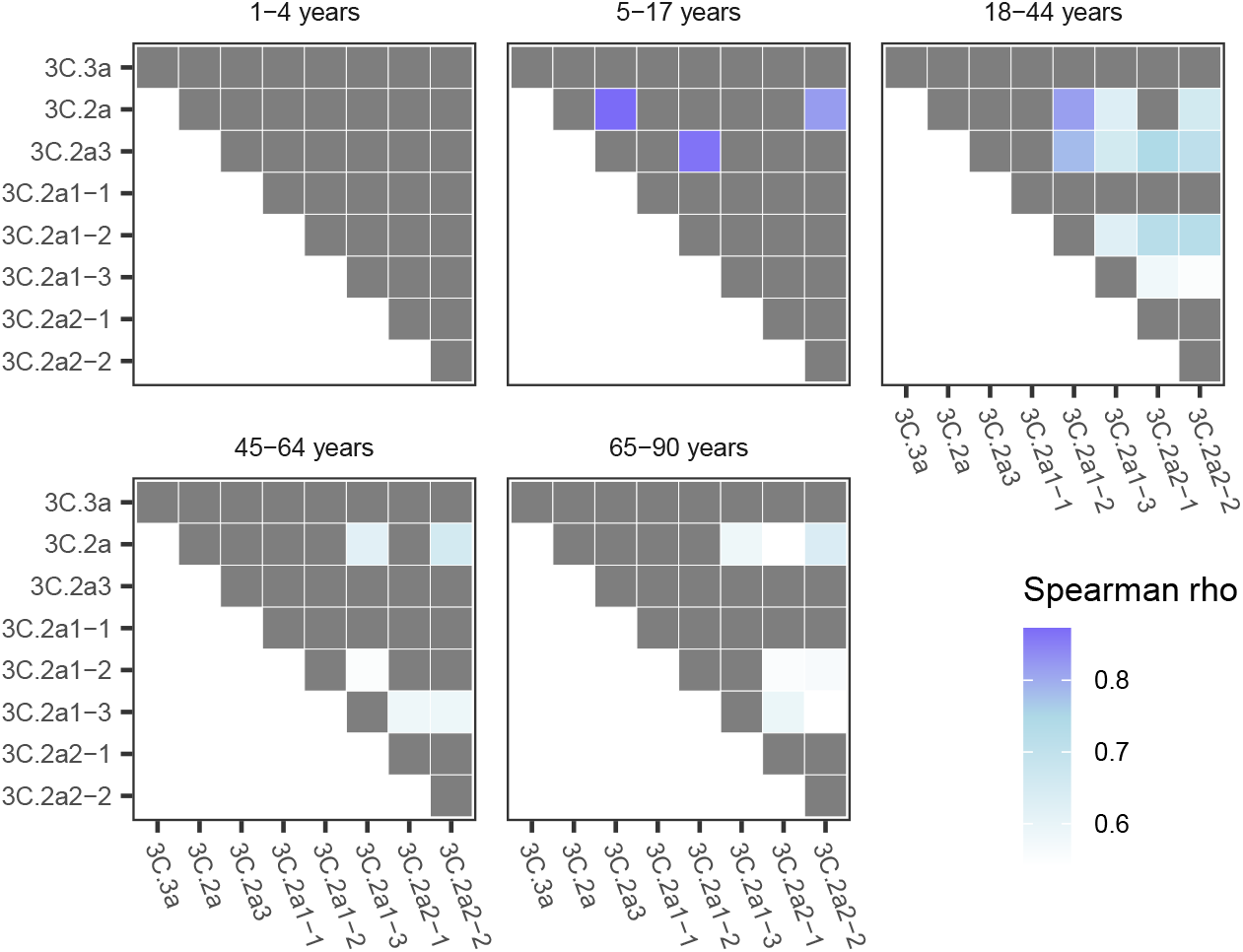
Compared to the youngest age group, older children and adults have significantly more weakly correlated titers to different strains. For each pair of viruses, we calculated the correlation in HA titers within each age group, averaging across 1000 random imputations of continuous titers. For each pair of strains, we used bootstrapping to test if the correlation for that pair in each age group was significantly weaker than in 1-4 year-olds. The cells are colored blue if the correlation is significantly weaker. Individuals with all undetectable titers were removed from these analyses.

**Fig S17.**
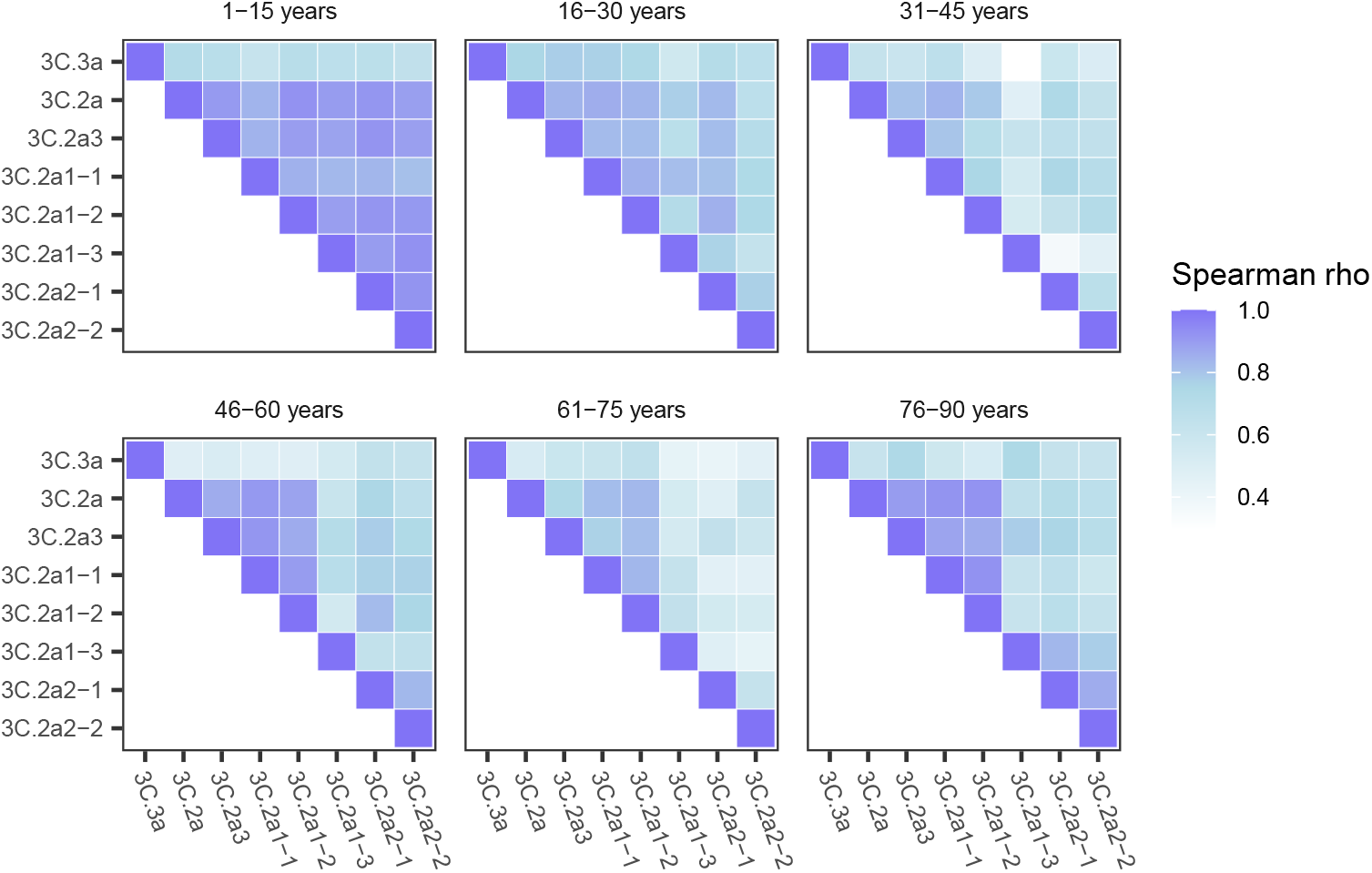
Weak correlations in adults’ HA titers between viral strains persist when age groups cover the same number of years. For these analyses, we randomly imputed continuous titer values between consecutive dilutions (e.g., a titer of 160 was replaced by a continuous value between 160 and 320 drawn with uniform probability). For each pair of viruses and each age group, we report the average Spearman correlation coefficient across 1000 replicate imputations. We removed individuals with all undetectable titers from this analysis.

**Fig S18.**
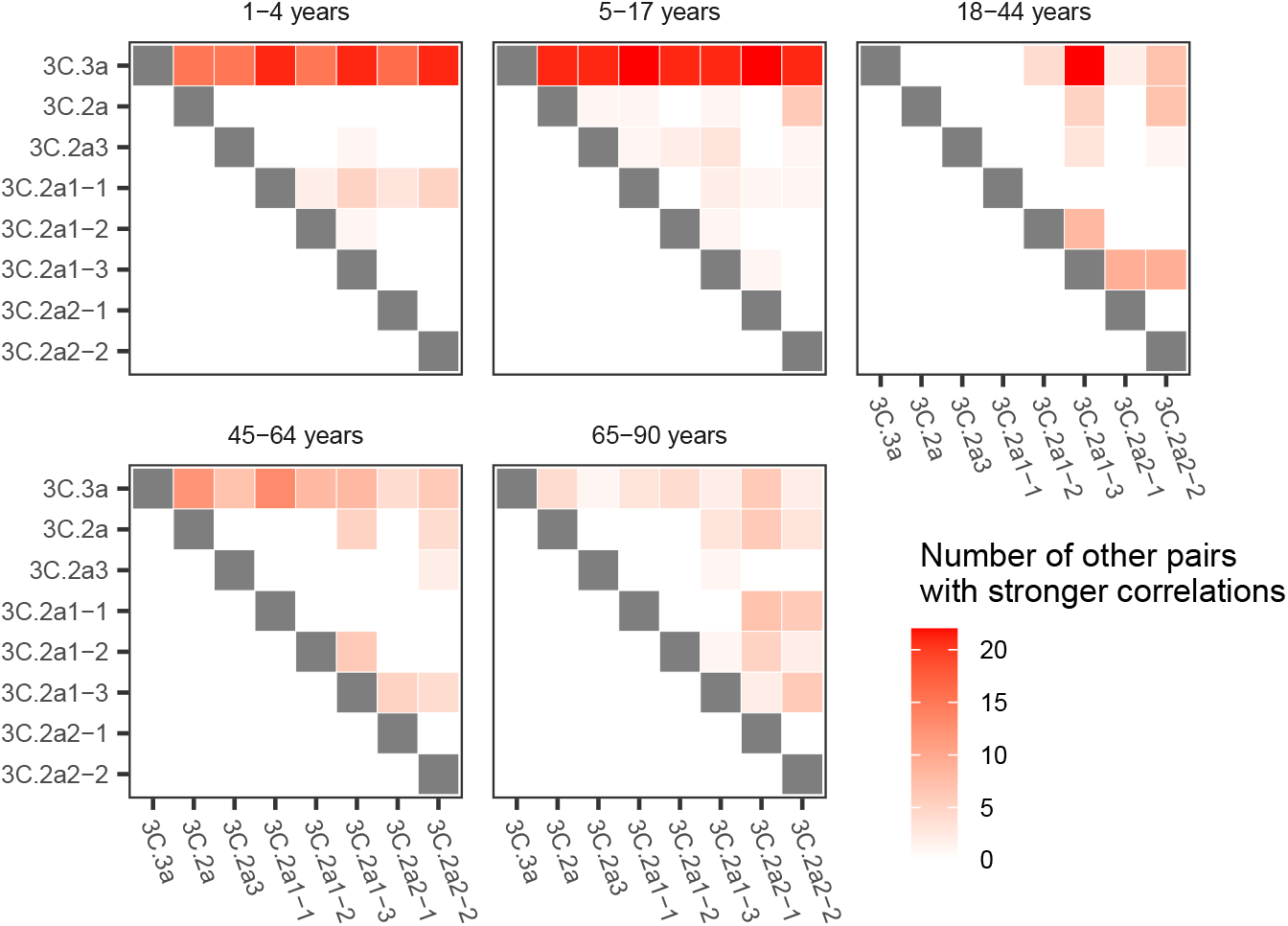
Titers to the 3C.3a virus had the weakest correlation with titers to other viruses. For each pair of viruses, we computed the correlation in HA titers in each age group, averaging across 1000 random imputations of continuous titer values. Within each age group, we used bootstrapping to compare the correlations across different pairs of strains. For each pair of strains, we counted how many other pairs of strains had significantly stronger correlations in the same age group. Strain pairs with weaker correlation than most other pairs appear in dark red. Individuals with all undetectable titers were removed from these analyses. Gray cells indicate the diagonal, where tests were not performed because the correlation is always 1

